# A Unified and Interpretable Geometric Optimization Framework for Intelligent Automated Implant Planning in CBCT from Single to Consecutive Multiple Missing Teeth

**DOI:** 10.64898/2026.10.03.26364640

**Authors:** Zilong Zeng, Yufan Jiang, Jie Li, Han Cao, Yuanwen Lu, Huilin Zhu, Junhao Xiong, Zhihui Tang, Narisu Hu, Hongyang Ma

**Affiliations:** School of Medical and Information Engineering, Gannan Medical University, Ganzhou, China; Department of Oral and Maxillofacial Surgery, Second Affiliated Hospital of Harbin Medical University, Harbin, China; School of Dentistry, Centre for Orofacial Regeneration, Rehabilitation and Reconstruction (COR3), University of Queensland, Brisbane, Australia; Light Cone IoT Technology (Shenzhen) Co., Ltd., Shenzhen, China; Second Clinical Division, Peking University School and Hospital of Stomatology, Beijing, China; Joint Laboratory of Artificial Intelligence in Oral Science, Binhai Chronic Disease Prevention and Control & Health Innovation Research Institute, Tianjin, China

**Keywords:** Dental implant planning, Cone-beam computed tomography (CBCT), Consecutive multiple missing teeth, Digital dentistry

## Abstract

Automated dental implant planning using artificial intelligence (AI) faces challenges in interpretability, safety assurance, and applicability beyond single-tooth cases. This paper presents a unified explainable framework for cone-beam computed tomography (CBCT) that handles both single-tooth and consecutive multiple missing teeth. The method combines nnU-Net-based anatomical segmentation with a transparent geometric optimization pipeline. Implant placement is formulated as a lexicographic multi-objective constrained optimization problem that enforces clinical safety margins while balancing bone engagement, mesiodistal spacing, and buccolingual position. For multi-tooth cases, a global coordination strategy ensures inter-implant parallelism, minimum spacing, and a common insertion path. The framework was validated on 20 single-tooth and 9 consecutive multi-tooth cases. The AI plans achieved a centroid distance of 1.60±1.05 mm for single-tooth cases and 1.22±0.32 mm for multi-tooth cases, with approximately 7° axial angle difference. All plans achieved 100% compliance with predefined safety thresholds. In a Turing-style test, experts were unable to distinguish AI from human intelligence (HI) plans under blinded conditions. By decoupling perception from reasoning and embedding clinical knowledge as explicit constraints, the framework offers an interpretable and clinically trustworthy solution for computer-aided implant dentistry.

## 1. Introduction

Dental implant therapy has become the gold standard for the rehabilitation of missing teeth, offering significant improvements in both oral function and aesthetic outcomes. The widespread adoption of cone-beam computed tomography (CBCT) has further enhanced pre-surgical planning by providing high-resolution three-dimensional visualization of maxillofacial anatomy, enabling clinicians to assess bone morphology, vital structures, and prosthetic constraints with unprecedented precision[1]. Despite these technological advances, traditional implant planning remains a predominantly manual and experience-driven process, which is not only time-consuming but also inherently susceptible to inter-clinician variability. This limitation becomes particularly pronounced in complex cases involving consecutive multiple missing teeth, where the need for coordinated implant placement, ensuring parallelism, optimal spacing, and aesthetic consistency across edentulous spans, introduces additional geometric and clinical challenges[2]. Consequently, there is a growing imperative to develop automated systems that can standardize and streamline implant planning while maintaining the rigor and transparency required for clinical adoption [3].

Recent advances in AI have demonstrated significant progress in medical image analysis[4-7], and implant planning has emerged as a promising application area[8-17]. Existing deep learning-based approaches generally fall into two categories: direct regression of implant parameters from CBCT volumes using convolutional neural networks[11-13, 16], and segmentation-based methods that treat the implant as a label to be predicted[10]. While these end-to-end models can achieve high accuracy on benchmark datasets, they suffer from fundamental limitations that hinder their clinical translation. First, they operate as black-box systems, offering little to no interpretability, a critical barrier in high-stakes surgical applications where clinicians must verify and assume responsibility for the final plan. Second, these models cannot explicitly enforce non-negotiable clinical safety constraints, such as minimum distances to the mandibular nerve canal, adjacent teeth, and cortical bone boundaries; instead, they rely on loss function penalties that do not guarantee constraint satisfaction at inference time. Third, most existing methods are designed exclusively for single-tooth scenarios and fail to generalize to the more complex multi-tooth cases that constitute a substantial portion of clinical practice. These three limitations highlight the need for a planning paradigm that combines the pattern recognition strength of AI with the transparency and controllability of rule-based reasoning[18].

In this paper, we propose a unified and explainable framework for automated implant planning that generalizes from single-tooth to consecutive multiple missing teeth. Our core idea is to digitize the expert clinician, emulating the iterative adjustment process that an experienced surgeon performs when finding an implant position. This process begins with a rough initial placement, using the midpoint between adjacent teeth and an axis interpolated from their long axes, followed by stepwise refinement to balance distances to the buccal and lingual bone walls, to adjacent teeth, and to neighboring implants when multiple implants are involved. To achieve this, we introduce three key innovations. First, we formulate a unified mathematical framework that seamlessly handles both single-tooth and multi-tooth edentulous spans, incorporating a lexicographic ordering of clinical priorities. Second, drawing inspiration from first-person shooter video games such as Counter-Strike, we adopt ray casting, the technique used to determine bullet hits, to measure critical distances from the implant surface to the bone walls.

Rays are cast from the implant’s shoulder and lateral surface in clinically relevant directions. The distance to the outer cortical bone surface is recorded via a backward-tracking procedure that avoids mistaking internal voids for the true bone boundary, providing an efficient, geometry-based assessment of bone thickness and subcrestal depth. Third, we design a lexicographic optimization algorithm that simulates the clinician’s decision hierarchy. This algorithm first ensures safety, addressing bone engagement and nerve clearance, then balances mesio-distal spacing to adjacent teeth, and finally balances the bucco-lingual bone thickness. For multi-tooth cases, we introduce a global coordination strategy that treats neighboring implants as additional obstacles, iteratively enforcing parallelism and minimum spacing. The entire planning pipeline is fully interpretable; every decision, from arch curve extraction to the final position adjustment, corresponds to a geometric operation that can be visualized and understood. The deep learning component is confined to anatomical segmentation, for which large annotated datasets are readily available, while the planning logic itself is encoded as explicit constraints and heuristics, eliminating the need for costly expert-annotated implant plans.

We validate our framework on a clinical dataset comprising 20 single-tooth and 9 consecutive multiple missing teeth cases. Experimental results demonstrate high geometric accuracy, full compliance with safety constraints, and strong agreement with human intelligence (HI) implants. A Turing-style clinical test further shows that experts cannot reliably distinguish AI-generated plans from HI-planned plans. By casting the expert clinician as a digital twin and leveraging game-engine-inspired geometric reasoning, our work provides a scalable, interpretable, and clinically trustworthy solution for computer-aided implant dentistry. The remainder of this paper is organized as follows. Section 2 presents the detailed methodology. Section 3 reports the experimental results on both single-tooth and multi-tooth cases. Section 4 discusses the findings, limitations, and future directions. Section 5 concludes the paper.

## 2. Material and Methodology

The main framework of our method is presented in Fig. 1. The overall workflow begins with CBCT acquisition and segmentation (A), where raw scans are processed by an nnU-Net to segment the jawbones, teeth, and mandibular canal (detailed in Section 2.1). This is followed by dental arch modeling and ROI extraction (B), which involves extracting tooth centroids, applying weighted PCA to fit a U-shape arch curve, and establishing local orientation fields to guide the arch-based ROI extraction (detailed in Sections 2.2 and 2.3). Based on the anatomical model, implant initialization (C) is performed using a midpoint with a SLERP axis for single-tooth gaps and arc-length division with interpolated axes for consecutive multi-tooth gaps (detailed in Section 2.4). Finally, implant planning safety construction and problem formulation (D) define single-implant safety constraints and multi-implant coordination constraints, formulating the planning as a multi-objective constrained optimization problem solved via projection and balancing until convergence (detailed in Section 2.6).

**Fig. 1.**
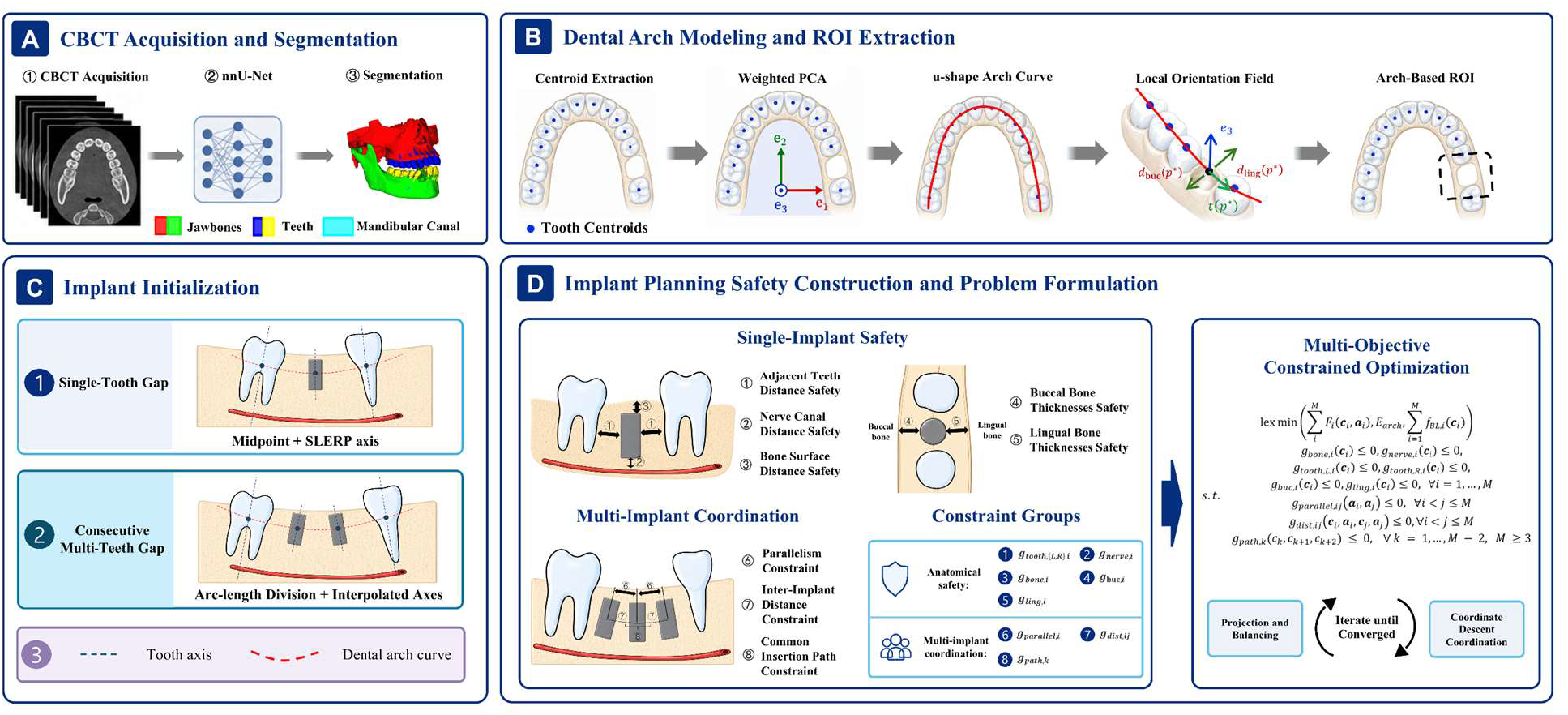
Main Framework.

Crucially, this optimization is strictly governed by four hard clinical safety constraints that must be satisfied before any objective optimization: (1) Bone Surface Distance Safety, ensuring a maximum subcrestal depth of 1.0 mm; (2) Nerve Canal Distance Safety, enforcing a minimum distance of 2.0 mm from the implant surface to the nerve canal in mandibular cases; (3) Adjacent Teeth Distance Safety, requiring a minimum interproximal distance of 1.5 mm to the mesial and distal teeth; and (4) Buccal and Lingual Bone Thickness Safety, demanding at least 2.0 mm of buccal bone thickness and 1.0 mm of lingual bone thickness. These constraints constitute a non-negotiable safety boundary [8]. Taking lexicographic precedence over the imbalance objectives, they rigorously enforce that the final implant position remains within the feasible anatomical region, preventing any violation of vital structures.

### 2.1. Data Acquisition and Preprocessing

#### 2.1.1. Clinical CBCT Dataset

This study employed a retrospective dataset of cone-beam computed tomography (CBCT) scans acquired from patients presenting with either single-tooth or consecutive multiple missing teeth (2–3 adjacent teeth). All CBCT scans were obtained from the database of Peking University Hospital of Stomatology. All images were reconstructed and stored in DICOM format. The study was approved by the Institutional Review Board of Peking University Hospital of Stomatology (IRB No. PKUSS202393156), and written informed consent was obtained from all patients. The details of inclusion criteria and ground truth of implant planning were described in Supplementary Materials SI-1 and SI-2.

#### 2.1.2. Data Preprocessing

To obtain accurate three-dimensional representations of relevant anatomical structures, we employed a pre-trained nnU-Net model [19] to segment each CBCT volume into five distinct labels: maxilla, mandible, maxillary teeth, mandibular teeth, and the mandibular nerve canal. After segmentation, individual tooth components were isolated using connected component analysis with a 6-connectivity for each jaw. This process separates individual teeth from the unified tooth label in the segmentation output, providing the necessary tooth-level information for dental arch modeling and implant planning.

### 2.2. Dental Arch Modeling

The dental arch provides a natural reference for determining the mesio-distal direction and the lingual/buccal orientation at any point along the jaw. We propose a weighted PCA-based method to extract the arch curve and its associated local direction fields.

#### 2.2.1. Teeth Centroids and Weighted PCA

Let *T* = {*T*_1_, *T*_2_, …, *T*_*N*_} denote the set of individual tooth components in the jaw (either maxillary or mandibular), obtained after connected-component filtering. For each tooth *T*_*i*_, its centroid ***m***_*i*_ ∈ ℝ^3^ is computed as the center of mass of its voxels, and a weight 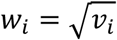 is assigned, where (*v*_*i*_) is the number of voxels of that tooth. The weights give larger teeth a stronger influence on the arch shape, which is clinically justified because posterior teeth are larger and contribute more to the overall arch form.

All centroids are collected in a matrix **M** ∈ ℝ^*N*×3^. The weighted mean and the weighted covariance matrix are 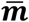 and ***C***, respectively:

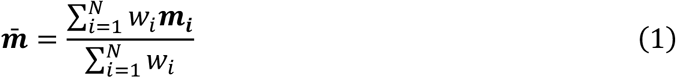

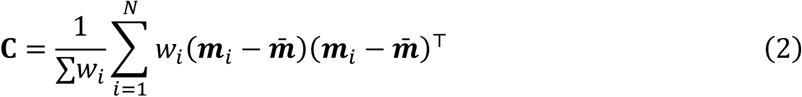

Eigen decomposition of **C** yields eigenvalues λ ≥ λ_2_ ≥ λ_3_ and corresponding eigenvectors **e**, **e**_2_, **e**_3_. The first two eigenvectors span the arch plane (the plane of the dental arch), with **e** representing the direction of maximum variance (roughly the mesio-distal direction) and **e**_2_ representing the bucco-lingual direction. The third eigenvector **e**_3_ is orthogonal to the arch plane and approximates the occlusal-apical direction.

#### 2.2.2. Arch Curve Fitting

The centroids are projected onto the arch plane using the orthonormal basis defined in Section 2.2.1:

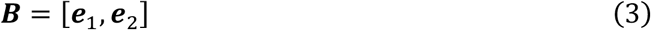

The projected coordinates are:

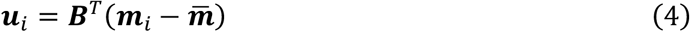

The projected points are sorted along the first coordinate, corresponding to the arch width direction. Curve fitting is then performed in this two-dimensional plane.

To reflect the anatomical morphology of the dental arch, we fit the projected centroids with a U-shape-constrained low-order polynomial. The polynomial degree is selected according to the number of available teeth, with a maximum degree of four. Before fitting, the width coordinate is normalized to the range from −1 to 1. The curve is estimated by weighted robust least squares using the tooth-size weights defined above.

The fitting objective includes a data fidelity term and a curvature-consistency regularization term. The latter penalizes curvature reversal relative to the dominant U-shaped bending direction, thereby reducing anatomically implausible local inflections caused by segmentation noise, missing-tooth gaps, or endpoint variation. After fitting, the curve is uniformly sampled and back-projected to the original three-dimensional coordinate system:

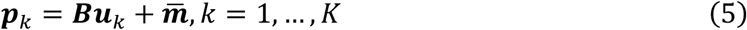

The final dental arch curve is denoted as:

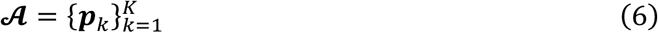

It is subsequently used to compute local tangent and buccolingual direction fields along the jaw.

#### 2.2.3. Local Direction Fields

For any point ***p*** on the arch curve, we compute the tangent vector ***t***(***p***) by finite differences (forward/backward/central depending on position) and project it onto the arch plane. The buccal and lingual direction vectors are defined as the outward and inward normals to the arch. Since the arch is approximately U-shaped, the inward direction points toward the center of the arch (i.e., toward the mean centroid 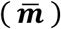. Specifically, we compute a candidate normal as ***n***(***p***) = ***t***(***p***) × **e**_3_ (with (**e**_3_) being the occlusal direction). The sign of ***n***(***p***) is adjusted so that it points toward the interior of the arch by comparing its dot product with the vector 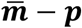. The final lingual direction is

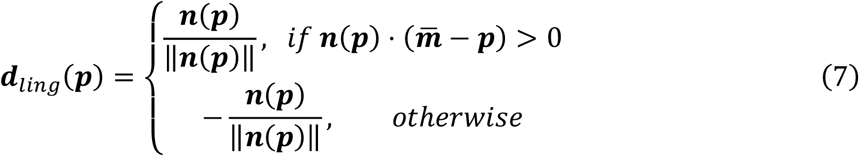

The buccal direction is simply ***d***_*buc*_(***p***) = −***d***_*ling*_(***p***). These direction fields are stored at a subset of arch points (sampled every 5 points along the curve) for later use.

### 2.3. Anatomy-Guided ROI Extraction

For a given edentulous site, we automatically extract a region of interest (ROI) that contains all relevant anatomy while minimizing the volume for subsequent optimization. The ROI is defined as a 3D bounding box oriented with the local arch coordinate system. Prior to ROI extraction, the edentulous span must first be identified from the segmented tooth set.

#### 2.3.1. Edentulous Span Detection

To automatically locate edentulous spans without training additional networks, we propose a purely geometric method based on dental arch arc-length. For each segmented tooth *T*_*i*_, we estimate its mesiodistal crown width *w*_*i*_ by performing PCA on its voxels: the second principal component (the first being the tooth’s long axis, used in Section 2.4.1) defines the mesiodistal direction, and the projection range of all voxels onto this axis gives *w*_*i*_. Each tooth centroid ***m***_*i*_ is then projected onto the arch curve ***A*** (Section 2.2.2) to obtain an arc-length coordinate *s*_*i*_. The sorted set {*s*_*i*_} yields gaps between adjacent teeth:

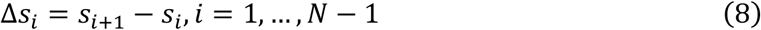

For each adjacent pair *T*_*i*_ and *T*_*i*+1_, the expected gap is estimated from their crown widths:

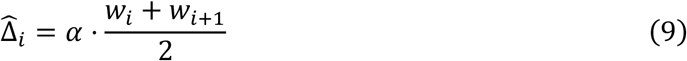

where *α* = 1.2 accounts for physiological interdental clearance. If 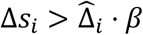, with *β* = 1.6, the span is flagged as edentulous, with *T*_*i*_ and *T*_*i*+1_ as its mesial and distal boundaries. The output is a list of edentulous spans, each defined by its boundary teeth, which directly feed into the ROI extraction described below.

#### 2.3.2. Single-Tooth Gap

Let the adjacent teeth labels be *L* (left) and *R* (right). Their voxel sets are denoted by *T*_L_ and *T*_R_. The centroid of the missing site is estimated as the midpoint of the two tooth centroids:

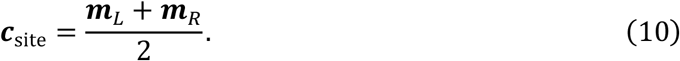

We locate the nearest point ***p***^*^ on the arch curve ***A*** *to* **c**_site_. The local basis at ***p***^*^ is given by {***t***(***p***^*^), ***d***_*buc*_(***p***^*^), **e**_3_}(or equivalently ***t, d***_*ling*_, **e**_3_).

The ROI is constructed as an axis-aligned box in this local coordinate system:

1. **Mesio-distal extent:** The projection of the two teeth onto the tangent direction ***t*** determines the minimum and maximum coordinates along this axis. A symmetric margin (e.g., 5 mm) is added beyond the tooth projections to ensure inclusion of the edentulous span.
2. **Bucco-lingual extent:** The projection of the teeth and surrounding bone onto the buccal direction *d*_*buc*_ is computed. Clinical safety margins (e.g., 2 mm beyond the outermost tooth points) are added to ensure that the full alveolar ridge is captured.
3. **Apico-coronal extent:** The entire height of the jaw at this location is included, from the lowest point of the jaw (or the mandibular canal, if present) to a level above the alveolar crest. Specifically, we search for voxels of the jaw bone within a region around the site and record the minimum and maximum *Z* coordinates (in the original image axes) after transforming to the local occlusal plane. Margins of 2–3 mm are added to cover the crest and basal bone.

The resulting box in the local coordinate system is transformed back to the original image coordinates using the affine transformation that maps local coordinates to world coordinates (based on ***p***^*^ and the local basis). The ROI volume is then cropped from the original segmentation image, and its affine matrix is updated accordingly to reflect the new origin. This process ensures that the ROI is anatomically aligned and contains all structures needed for planning.

#### 2.3.3. Extension to Consecutive Multiple Missing Teeth

For a consecutive edentulous span of length *M* (e.g., missing teeth (*T*_*L*_, *T*_*L*+1_, …, *T*_*R*_), we consider the two boundary teeth (the most mesial and distal remaining teeth) as the anchors. The ROI is defined in the same way, but the centroid of the missing site is taken as the midpoint between these two anchor teeth. The local basis is obtained from the arch point nearest to that midpoint. The mesio-distal extent must be expanded to cover the entire gap, so we compute the projection of the two anchor teeth and then extend the box by half the estimated gap length on each side, ensuring that the entire edentulous span is enclosed. If intermediate teeth are partially present (e.g., root remnants), they are also included automatically as they lie within the bounding box.

### 2.4. Implant Initialization

#### 2.4.1. Axis Initialization

For each adjacent tooth (or for the two boundary teeth in a multi-tooth gap), we compute its long axis using PCA on its voxel coordinates. Let ***a***_*L*_ and ***a***_*R*_ be the unit vectors representing the axes of the left and right tooth (pointing from apex to crown). The initial implant axis ***a***_0_ is obtained by spherical linear interpolation (SLERP) with parameter *t* = 0.5:

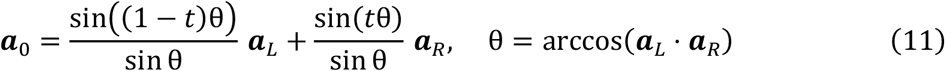

This yields a direction that smoothly varies between the two adjacent axes, mimicking the natural alignment of a prosthetic crown. The vector is then normalized. For a single missing tooth, this provides a physiologically plausible starting point.

#### 2.4.2. Position Initialization

For single missing tooth, the initial implant center ***c***_O_ is set as the midpoint of the centroids of the two adjacent teeth:

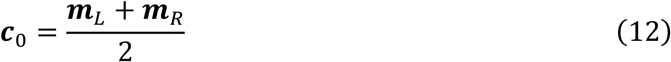

This places the implant in the center of the edentulous span in the mesio-distal direction. For consecutive multiple missing teeth, the two remaining teeth at the mesial and distal ends of the edentulous span are used as boundary anchors. The initial implant centers are evenly distributed between these two anchors along the dental arch.

For *M* planned implant positions, the target locations are assigned according to the fractions 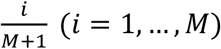. If the dental arch segment is unavailable, the same fractions are applied by linear interpolation between the two boundary tooth centroids. These initial centers are subsequently refined by the optimization procedure (detailed in Section 2.6). For most patients with consecutive multiple missing teeth included in this study, the number of missing teeth is two. For patients with more than two missing teeth, clinically, implants are typically placed only at the two ends of the edentulous span to perform bridge restoration. Therefore, for such patients, although M implants are retained during both the initialization and optimization phases (Fig. S5), the final outputs ultimately retain only the two implants at the ends of the edentulous span. (Fig. S4).

#### 2.4.3. Implant Size Selection

The implant diameter and length are automatically selected based on tooth-specific preferences and local bone geometry. The edentulous span (Section 2.3.1) is mapped to an FDI number via the sorted connected component sequence (Section 2.1.2). The preferred diameter is 3.3 mm for incisors and canines, 4.1 mm for premolars, and 4.8 mm for molars.

Buccolingual bone width *BW* is measured at the span midpoint using ray casting along the local buccal/lingual directions (Section 2.2.3) with forward-backward tracking (Section 2.5.1) to identify cortical intersections. The diameter upper bound is *D*_*max*_ = *BW* − 3.0 *mm* (1.0 mm margin on the lingual side and 2.0 mm margin on the buccal side). The selected diameter is the preferred value if it does not exceed *D*_*max*_; otherwise, it is downgraded to the next smaller size from {3.3 mm, 4.1 mm, 4.8 mm} that satisfies the bound. If none satisfies the constraint, the case is flagged as infeasible.

Vertical bone height *BH* is measured at the same point along the initial implant axis (Section 2.4.1): from the crest to the mandibular canal (mandible) or to the sinus floor (maxilla). The maximum allowable length is *L*_*max*_ = *BH* − 1.0 − *D*_*safe*_, where *D*_*safe*_ is 2.0 mm for the mandible and 1.5 mm for the maxilla. The preferred length is 10 mm; if 10 mm exceeds *L*_*max*_, 8 mm is selected. If 8 mm also exceeds *L*_*max*_, the case is flagged as infeasible.

For consecutive multi-tooth cases, each implant is sized independently. For spans longer than a single tooth, *BW* and *BH* are measured at each candidate position, and the minimum values are used to ensure safety.

### 2.5. Distance Measurement Methods

Accurate distance measurement is required to optimize implant placement relative to surrounding anatomy. We use two complementary strategies: ray casting with backward endpoint refinement for bone surface distances, and Euclidean distance transform for distances to discrete anatomical structures, including adjacent teeth and the mandibular canal. All distances are computed in millimeters using the ROI voxel spacing and affine transformation.

#### 2.5.1. Ray Casting for Bone Surface Distances

The proposed bone surface distance measurement is inspired by ray-casting and hit-detection mechanisms in game engines[20, 21]. As illustrated in Fig. 2A, shooting games emit a ray from a viewpoint or weapon along a specified direction to identify the hit point on a target. Fig. 2B shows the same principle in 3D space, where a ray origin, ray path, and impact point define a directional collision query. In our framework, this idea is adapted to implant planning by casting rays from implant surface sampling points toward anatomically meaningful directions and detecting their intersections with the outer jaw surface.

**Fig. 2.**
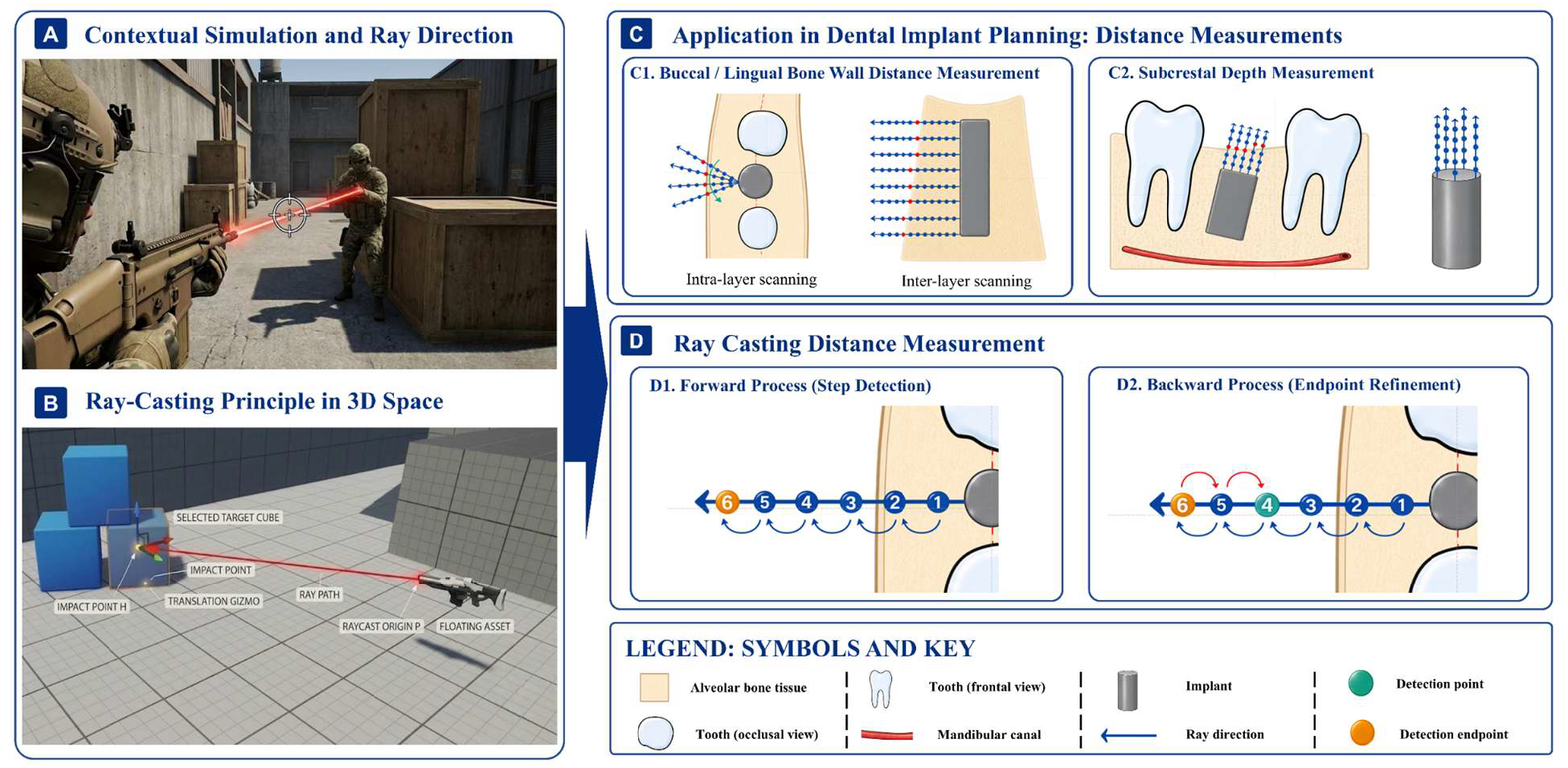
Ray Casting Distance Measurement.

Given a starting point on the implant surface, a ray direction, and a binary jaw mask, points are sampled along the ray at fixed step intervals. Each sampled point is converted into ROI voxel coordinates, and its inside/outside state with respect to the jaw mask is recorded together with its physical distance from the starting point. The ray directions are determined according to the measurement task. For buccal and lingual bone wall assessment, the local buccal and lingual direction fields derived from the dental arch model in Section 2.2.3 are used as anatomical references. Specifically, the direction field nearest to the current implant center provides the initial buccal or lingual orientation, which is then aligned with the local geometry defined by the adjacent teeth and the implant axis. A small fan of rays centered on this direction is emitted from the implant lateral surface at multiple axial levels (Fig. 2C1). This design allows the measurement to follow the patient-specific dental arch while remaining robust to local bone-wall irregularities. For platform-to-bone surface measurement, rays are emitted from points sampled on the implant platform circumference along the implant-axis-related direction (Fig. 2C2). The resulting distances estimate the platform embedment depth relative to the outer bone surface.

To robustly identify the true outer cortical boundary, each ray is processed using a forward and backward procedure. In the forward process (Fig. 2D1), the ray is propagated step by step from the implant surface to generate an ordered sequence of sampled points and jaw-mask states. However, the first label transition along the ray may correspond to an internal void, marrow space, or segmentation defect. Therefore, in the backward process (Fig. 2D2), the ray is first extended by a fixed distance of 50 mm from its initial point, thereby extending beyond the jaw, and the sampled sequence is traversed in reverse from the outermost end toward the implant. The outer bone surface is defined as the outermost jaw-label segment encountered during this reverse traversal. The distance from the implant sampling point to this refined endpoint is recorded as the bone surface distance.

#### 2.5.2. Distance Transform for Discrete Structures

For adjacent teeth and the mandibular canal, which are represented as discrete voxel masks, we precompute Euclidean distance transform maps. Given a binary mask ***S*** of the structure, we compute for every voxel its Euclidean distance to the nearest voxel of ***S***, using the physical voxel spacings to obtain distances in millimetres. This produces a distance map *D*_***S***_(**x**) for all voxel coordinates **x**.

For a given implant pose, we obtain the set of implants voxels *I*(***c, a***). The minimum distance from the implant to the structure is then:

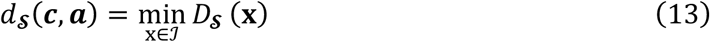

This is efficient because the distance map is computed once per ROI and reused for all implant positions during optimization. This method is applied to measure the adjacent teeth distances *d*_*L*_ and *d*_*R*_, and mandibular canal distance *d*_*nerve*_.

### 2.6. Multi-Objective Constrained Optimization

We formulate the implant placement problem as a lexicographic multi-objective optimization with explicit clinical safety constraints. Let ***c*** ∈ ℝ^3^ denote the implant center in physical coordinates (mm). The implant axis ***a*** (determined in Section 2.4.1) is kept fixed during the position optimization; the orientation is thus not part of the decision variables here.

#### 2.6.1. Problem Formulation for a Single Implant

Using the distance measurements defined in Section 2.5, we define the following functions (all depend on ***c***; the dependence on ***a*** is omitted for brevity):

1. **Bone surface distance safety** *g*_*bone*_(***c***):

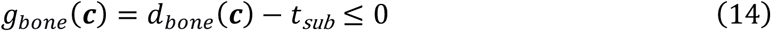

where *d*_*bone*_(***c***) is measured by ray casting from the implant shoulder (Section 2.5.1), and *t*_*sub*_ denotes the maximum allowable subcrestal depth, imposed as 1.0 mm.
2. **Nerve canal distance safety** (mandible only) *g*_*nerve*_(***c***):

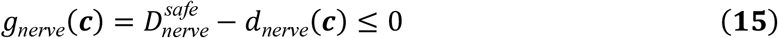

where *d*_*nerve*_(***c***) is obtained via distance transform of the nerve canal (Section 2.5.2), and 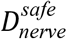 denotes the safety threshold, imposed as 2.0 mm.
3. **Adjacent teeth distance safety** *g*_*to L*_(***c***), *g*_*toothR*_(***c***):

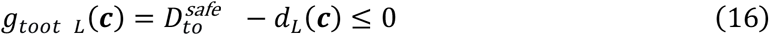

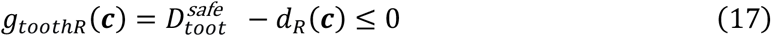

where *d*_*L*_(***c***), *d*_*R*_(***c***) are obtained via distance transform of the left and right teeth (Section 2.5.2), and 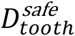 denotes the safety threshold, imposed as 1.5 mm.
4. **Buccal and lingual bone thicknesses safety** *g*_*buc*_(***c***), *g*_*ling*_(***c***):

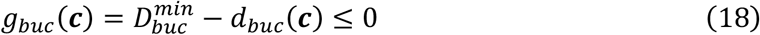

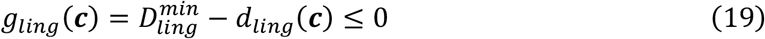

where *d*_*buc*_(***c***), *d*_*ling*_(***c***) are measured by ray casting from the implant lateral surface (Section 2.5.1, and 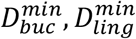 denotes the safety thresholds, imposed as 2.0 mm and 1.0 mm, respectively.

In addition to satisfying all safety constraints, we wish to balance the implant between the two adjacent teeth and between the buccal and lingual bone plates. Thus, two objective functions are given:

1. The first one is the mesio-distal imbalance function, which is used to balance the implant position along the mesio-distal direction:

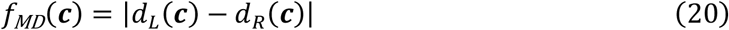
2. The second one is the bucco-lingual imbalance function, which used to balance the distance from implant surface to the bucco and lingual bone surface:

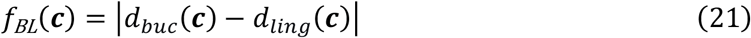

Clinically, ensuring safety is paramount; only after all constraints are satisfied should the balance between the mesio-distal and bucco-lingual directions be optimized. This natural priority suggests a lexicographic ordering of the objectives: first minimize *f*_*MD*_ (subject to all constraints), and then, among the optimal solutions for *f*_*MD*_, minimize *f*_*BL*_. Thus, the overall problem can be stated as:

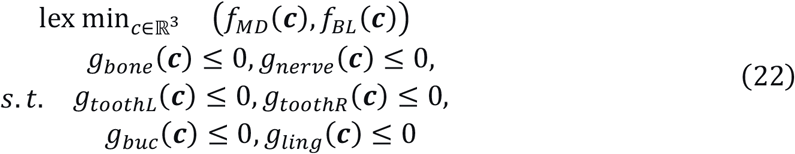

All functions are non-smooth (they involve minima of distances) and non-convex due to the complex geometry of bone and teeth. Consequently, classical gradient-based methods are not directly applicable.

#### 2.6.2. Extension to Consecutive Multiple Implants

For a gap of *M* consecutive missing teeth (*M* ≥ 2), we denote the set of implants as 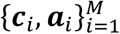, where ***c***_*i*_ ∈ ℝ^3^ is the center of the *i*^*th*^ implant and ***a***_*i*_ (unit vector) is its axis. The individual safety constraints and balance objectives defined in Section 2.6.1 are extended to each implant by appending the subscript *i* (e.g., *g*_*bone,i*_(***c***_*i*_), *f*_*BL,i*_(***c***_*i*_)). In addition to these individual terms, multiple inter-implant constraints must be enforced to ensure prosthetic feasibility and long-term stability.

##### 1) Parallelism constraint

The axes of any two implants should be approximately parallel to allow a common prosthetic restoration and to avoid unfavorable stress distribution. The angular deviation between ***a***_*i*_ and ***a***_*j*_ is bounded by a clinical threshold *θ*_max_ (e.g., 5°):

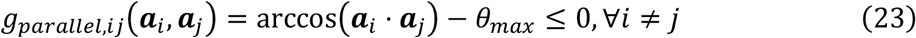

This constraint is enforced for every pair of implants in the edentulous span.

##### 2) Inter-implant distance constraint

To preserve bone integrity and avoid implant-to-implant contact, the surface clearance between the two implants is constrained to be no less than a prescribed safety margin. Each implant is represented by a cylindrical model with center, unit axis, and radius. Let ***d*** = ***c*** − ***c***_*i*_ denote the center-to-center displacement in physical coordinates and let ***ā*** denote the normalized mean axis. The displacement component orthogonal to the mean axis and the estimated surface clearance are defined as:

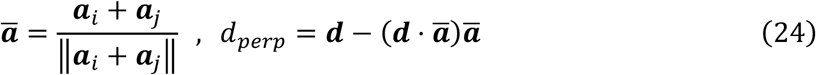

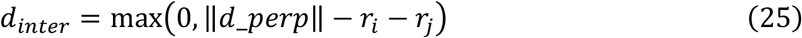

The constraint is:

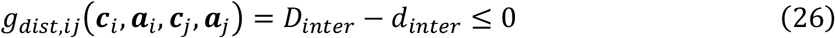

where *D*_*inter*_ is the minimum required surface clearance (e.g., 3.0 mm). When the estimated surface clearance falls below this margin, the implant centers are moved symmetrically apart along the local arch tangent if available; otherwise, they are separated along the line connecting the two centers.

##### 3) Common insertion path constraint

For cases involving three or more consecutive missing teeth, the implants must share a common insertion path (also called a common trajectory) to enable a screw-retained fixed prosthesis. This condition requires that the centers of the implants lie approximately on a straight line when projected onto the occlusal plane, i.e., the vector connecting adjacent implant centers should be nearly collinear.

Let ***p***_*i*_ denote the projection of **c**_*i*_onto the occlusal plane (or equivalently onto the arch plane defined in Section 2.2). For a sequence of implants ordered mesiodistally, define the direction vectors between consecutive projected centers:

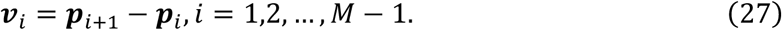

The common insertion path constraint requires that the angle between any two consecutive vectors **v**_*i*_and **v**_*i*+1_be below a threshold *ϕ*_*max*_(e.g., 5°):

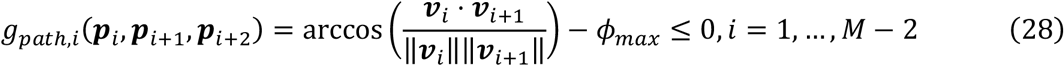

For *M* = 2 (two implants), a common insertion path is automatically satisfied if the two axes are parallel (the parallelism constraint already ensures that the two centers can be connected by a straight line without violating the prosthesis design). Therefore, this constraint is only active when *M* ≥ 3.

##### 4) Arch-distribution prior

For multiple consecutive missing teeth, the mesiodistal term is not formed by independently balancing each implant against the same two boundary teeth. Instead, it is represented by an arch-distribution prior over all implant centers. Let *L* denote the arc length of the dental arch segment between the two boundary teeth, and let *s*(***c***_*i*_) denote the arc-length coordinate of the projection of the center of the *i*^*th*^ implant onto this segment. The arch-distribution prior encourages uniform spacing of the implants along the dental arch:

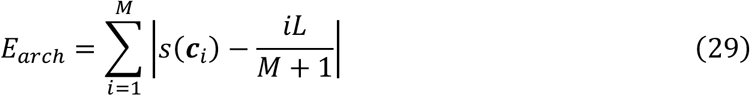

where *iL*/(*M* + 1) represents the desired uniformly distributed position of the *i*^*th*^ implant. When *M* = 2, the target positions reduce to *L*/3 and 2*L*/3. The arch-distribution term is a soft prior and does not override safety or inter-implant constraints.

##### 5) Optimization formulation

Combining the individual constraints defined in Section 2.6.1, the inter-implant constraints defined above, the arch-distribution prior, the complete optimization problem for *M* ≥ 1 implants can be uniformly formulated as:

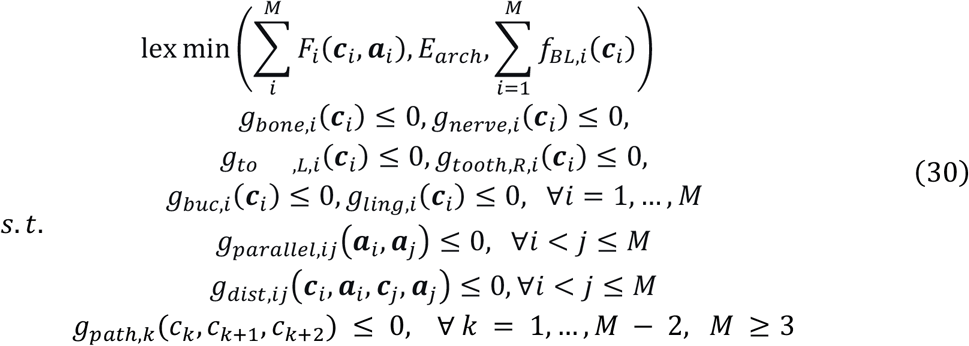

where *F*_*i*_(***c***_*i*_, ***a***_*i*_) aggregates the individual safety constraint variations for the *i*^*th*^ implant:

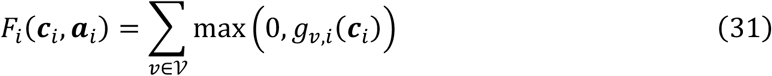

with *V* = {*bone, nerve, toothL, toothR, buc, ling*} (note that *g*_*nerve,i*_ is only active for mandibular cases).

The first lexicographic level minimizes the total individual safety violations. The second level minimizes *E_arch_*, which encourages uniform distribution of implants along the dental arch. The third level minimizes the sum of buccolingual imbalance terms. When *M* = 1, the inter-implant constraints are automatically inactive, and *E*_*arch*_ reduces to |*s*(*c*_1_) − *L*/2|, which is equivalent to the mesiodistal balance objective *f_MD_*(***c***_1_) of Section 2.6.1. Thus, the single-implant problem is a special case of this unified framework.

#### 2.6.3. Solution Algorithm

We solve the optimization problems formulated in Sections 2.6.1 and 2.6.2 using an iterative algorithm that alternates between projecting onto the feasible sets of the constraints and taking steps that reduce the imbalances. The algorithm exploits clinically meaningful directions: the implant axis ***a***, the arch tangent *t*(***p***) (obtained from the dental arch model), and the buccal/lingual directions ***d***_*buc*_(***p***), ***d***_*ling*_(***p***). These directions remain fixed during the optimization for a given edentulous site.

Let *P*_*g*_ denote a projection operator that moves **c** along a specific direction to reduce the violation of a constraint *g*(**c**) ≤ 0. Because the constraints are not convex sets, the projection is heuristic: we move in the direction that is known to alleviate the violation (e.g., along the axis to adjust bone depth, or occlusally to increase nerve clearance). Similarly, for the imbalance objectives, we define descent directions ***d***_*MD*_ and ***d***_BL_ that are expected to decrease *f*_*MD*_ and *f*_*BL*_ respectively.

**Algorithm 1** (Lexicographic optimization via sequential projection and balancing)

1. Initialize ***c***^**0**^ as the midpoint of the adjacent tooth centroids.
2. Repeat for *k* = 0,1,2, … until convergence (‖***c***^k+1^ − ***c***^*k*^ ‖ < *ε*):
  - **Bone depth projection:** If *g*_*bone*_(***c***^*k*^) > 0, move ***c***^*k*^ along −***a*** (maxilla) or +***a*** (mandible) by a step *δ_bone_* to reduce *g_bone_*. This step aims to satisfy *g_bone_*(***c***^k+1^) ≤ 0.
  - **Nerve safety projection (mandible only):** If *g_nerve_*(***c***^*k*^) > 0, move ***c***^*k*^ along +***a*** occlusally by *δ*_*nerve*_.
  - **Mesio-distal balancing:** Compute the signed imbalance Δ_*MD*_ = *d_L_*(***c***^***k***^) − *d_R_*(***c***^***k***^). The descent direction ***d***_*M*_ is the arch tangent ***t***(***p***^*^) with sign chosen to reduce Δ_*MD*_ (i.e., if Δ_*MD*_ > 0, move toward the left tooth; if Δ_*MD*_ < 0, move toward the right). Update ***c***^***k***+**1**^ = ***c***^***k***^ + *γ*_*MD*_***d***_*M*_, where *γ*_MD_ is a step size chosen to avoid violating any safety constraint.
  - **Bucco-lingual balancing:** Compute Δ_*BL*_ = *d_buc_*(***c***^***k***^) − *d_ling_*(***c***^***k***^). The descent direction ***d***_*BL*_ is the buccal direction ***d***_*buc*_(***p***^*^) ifΔ_*BL*_ > 0 (move buccally to reduce buccal distance), or the lingual direction ***d***_*ling*_(***p***^*^) ifΔ_*BL*_ < 0. Update ***c***^***k***+**1**^ = ***c***^***k***^ + *γ*_*BL*_***d***_*BL*_ with a suitable step size.
3. Output the final implant position ***c***^*^. The step sizes δ and γ are adaptively chosen based on the current violation or imbalance, and are reduced if a move would cause a constraint violation. This ensures that the algorithm remains within the feasible region once it is reached.

For *M* ≥ 2, the algorithm is extended by embedding the single-implant refinement within an outer loop that also enforces inter-implant constraints. At each outer iteration, the following steps are performed:

1. **Individual refinement**: each implant ***c***_*i*_ is independently refined using Algorithm 1, treating the other implants as fixed obstacles.
2. **Parallelism correction**: if the angular deviation between any pair of axes ***a***_*i*_ and ***a*** exceeds *θ*_*max*_, the two axes are iteratively aligned toward their mean direction using spherical linear interpolation:

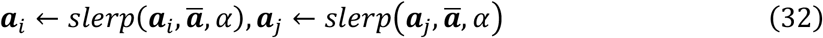

where ***ā*** = (***a***_*i*_ + ***a***_*j*_)/Il***a***_*i*_ + ***a***_*j*_Il and *α* is a step size (e.g., 0.5). This process is repeated until the parallelism constraint is satisfied.
3. **Distance correction**: If the surface clearance between two implants falls below *D*_inter_, the two centers are moved symmetrically apart. The correction follows the local arch tangent in the transverse plane when dental-arch metadata are available; otherwise, it follows the line connecting the two centers.
4. **Common path correction** (for *M* ≥ 3): If the common insertion path constraint is violated, the misaligned center ***p***_***k***+**1**_ is projected onto the line defined by ***p***_*k*_ and ***p*** _*k*+2_, and ***c***_*k*+1_ is updated accordingly.
5. **Arch-distribution prior**: A soft correction is applied that nudges each implant toward its target position *iL*/(*M* + 1) along the dental arch segment.

These steps are repeated until all constraints are satisfied and the implant positions converge (typically within 3–5 outer iterations). This approach effectively decouples the multi-implant problem into a sequence of single-implant subproblems, while maintaining global consistency through the correction steps.

### 2.7. Evaluation Framework

To assess the proposed framework, we evaluated AI plans from three aspects: geometric accuracy, clinical safety verification, and expert subjective assessment. All evaluations were compared against the ground truth defined by HI implants.

#### 2.7.1. Matching AI and HI Implants

For each planned implant (AI or HI), we extracted its position, axis, and endpoints for comparison. The implant axis was estimated using PCA on the implant voxels. The shoulder (top) and apex (bottom) were defined as the extreme points along this axis. For multi-implant cases, AI and HI implants were matched by their mesio-distal order along the dental arch.

#### 2.7.2. Single-Implant Geometric Accuracy

For each matched implant pair, we computed seven metrics to quantify how closely the AI implant matches the HI implant (Fig. 3A). The centroid distance (CD, mm) measures the Euclidean distance between the volumetric centroids of the two implants. The shoulder deviation (ShD, mm) and apex deviation (AD, mm) quantify the distances between the corresponding endpoints of the two implants. The axial angle difference (AAD, °) is the acute angle between the two estimated implant axes. The Dice coefficient (DICE) measures the volumetric overlap ratio between the two implant masks. The average surface distance (ASD, mm) computes the mean distance from each point on the AI implant surface to the nearest point on the HI implant surface. Finally, the modified Hausdorff distance (MHD, mm) represents the 95th percentile of the bidirectional surface distances, providing a robust measure of maximum discrepancy. These metrics were computed for all implants and reported separately for the single-tooth and multi-tooth cohorts.

**Fig. 3.**
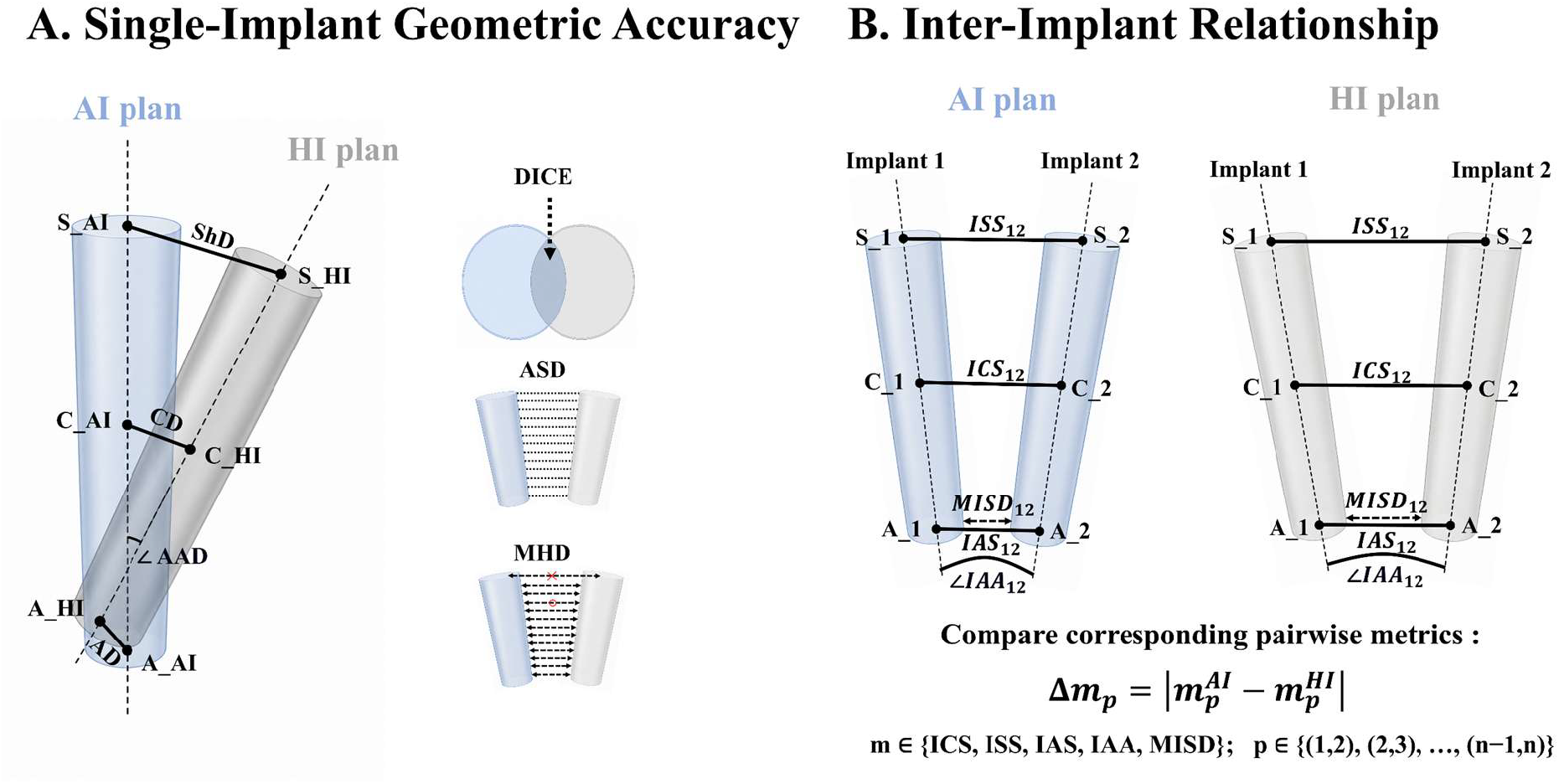
Illustration of the evaluation metrics. (A) Single-implant geometric accuracy metrics (ShD, CD, AD, AAD, DICE, ASD, MHD). (B) Inter-implant relationship metrics (ICS, ISS, IAS, IAA, MISD). Blue and gray models denote the AI and HI implant, respectively. Pairwise differences are computed as 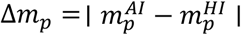.

#### 2.7.3. Inter-Implant Relationship

For cases with multiple consecutive missing teeth (*M* ≥ 2), single-implant accuracy alone does not guarantee prosthetic feasibility. We also evaluated the relative configuration between adjacent implants using five pairwise metrics (Fig. 3B). The inter-implant centroid spacing (ICS, mm) measures the Euclidean distance between the centroids of two adjacent implants. The inter-implant shoulder spacing (ISS, mm) and inter-implant apex spacing (IAS, mm) quantify the distances between the corresponding endpoints of two adjacent implants. The inter-implant axial angulation (IAA, °) is the angular deviation between the axes of two adjacent implants. The minimum inter-implant surface distance (MISD, mm) measures the minimum Euclidean distance between the surfaces of two adjacent implants, serving as a direct indicator of bone bridge integrity. For each metric, we calculated the absolute differences between AI and HI plans (Δ*ICS*, Δ*ISS*, Δ*IAS*, Δ*IAA*, Δ*MISD*) to quantify whether the AI framework preserves the expert-defined relative configuration. For spans with *M* > 2, these metrics were evaluated exclusively between the two retained implants at the ends of the edentulous span (Section 2.4.2).

#### 2.7.4. Clinical Safety Assessment

To evaluate clinical safety, for each implant, we measured the minimum distances (in mm) from the implant surface to adjacent mesial and distal teeth (≥1.5 mm), the buccal bone wall (≥2.0 mm), the lingual bone wall (≥1.0 mm), and, for mandibular cases, the inferior alveolar nerve canal (≥2.0 mm). These measurements verify whether the implant remains within safe anatomical margins. A plan was considered fully compliant only if all thresholds were simultaneously satisfied.

#### 2.7.5. Turing-style Clinical Test

In addition to the direct Likert scoring, we conducted a binary discrimination test (Turing-style) [22]. The experts were presented with a randomly interleaved set of AI-generated and HI-generated plans, without knowing the source. For each plan, they were forced to classify it as either “AI-generated” or “HI-generated.” The primary outcome of this test was the misclassification rate (i.e., the percentage of cases where the expert’s judgment was incorrect). A misclassification rate not significantly different from 50% (random guessing) would indicate that the AI plans are visually and clinically indistinguishable from HI plans.

#### 2.7.6. Expert Clinical Rating (ECR)

To evaluate clinical plausibility beyond objective geometric metrics, five expert clinicians (each with >10 years of clinical practice) independently reviewed all AI plans in a randomized and blinded fashion. Using a 5-point Likert scale, they rated each plan from 1 (clinically unacceptable, requiring major revision) to 5 (excellent, ready for surgery without modification). The scoring criteria were stratified by case complexity, with detailed rubrics provided in Supplementary Materials. Inter-rater reliability among the five experts was assessed using the intraclass correlation coefficient (ICC).

#### 2.7.7. Statistical Analysis

All continuous variables are reported as mean ± standard deviation (SD). For comparing AI-generated plans against HI-planned ground-truth, we employed the Wilcoxon signed-rank tests. A p-value of < 0.05 was considered statistically significant. All statistical analyses were performed using Python (SciPy v1.10).

## 3. Experimental results

This study enrolled 29 clinical cases, comprising 20 single-tooth and 9 consecutive multiple missing teeth cases. To validate the proposed algorithms, we performed a systematic analysis from three complementary angles: qualitative visual comparisons, objective geometric and safety metrics, and subjective expert clinical evaluations. The objective metrics were reported separately for the single and multiple missing-tooth groups. Subjective assessments, which included expert clinical scoring and a Turing-style test, were conducted on the combined dataset of all cases to boost statistical power.

### 3.1. Single-tooth Implant Planning

#### 3.1.1. Qualitative Analysis

At the 3D view (as shown in Fig 4), the AI plans were highly consistent with the HI plans across all subjects, with the long axes of the implants and the insertion paths being generally aligned, and no obvious deviations were observed. Further comparisons via multi-view 2D cross-sections (as shown in Fig 5) showed no significant differences between the AI and HI plans in key indicators. The safety distances to the nerve canal, the distance distribution between the implants and bone surfaces, and the distance with adjacent teeth were all very close to those of the HI plans, with no observed trend of displacement toward the adjacent teeth. The buccolingual bone volume allocation was reasonable in both groups, with only slight differences in local residual bone wall thickness and central alignment. Overall, the AI plans for single-tooth implants demonstrated good consistency with the HI plans.

**Fig. 4.**
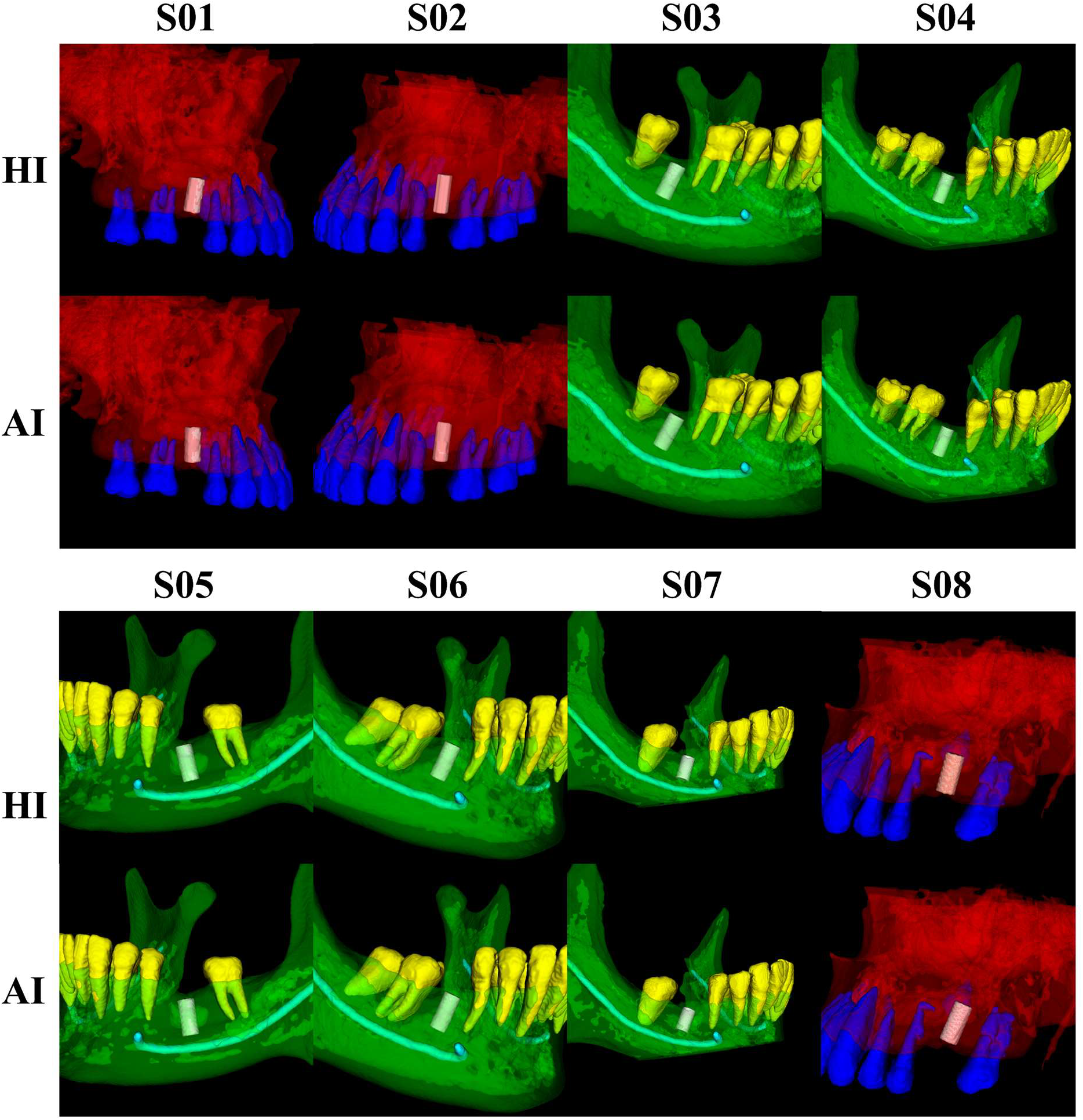
Comparison of AI and HI implant placement positions for eight subjects with single-tooth missing. For each subject, the planned implants are displayed within the anatomical context of the jawbone, adjacent teeth, and the mandibular canal, facilitating direct visual comparison between the two planning methods. Red, green, cyan, blue, yellow and white indicate the maxillary bone region, the mandibular bone region, the nerve canal, the maxillary teeth, the mandibular teeth, and the implant respectively. (HI means human intelligence, and AI means artificial intelligence)

**Fig. 5.**
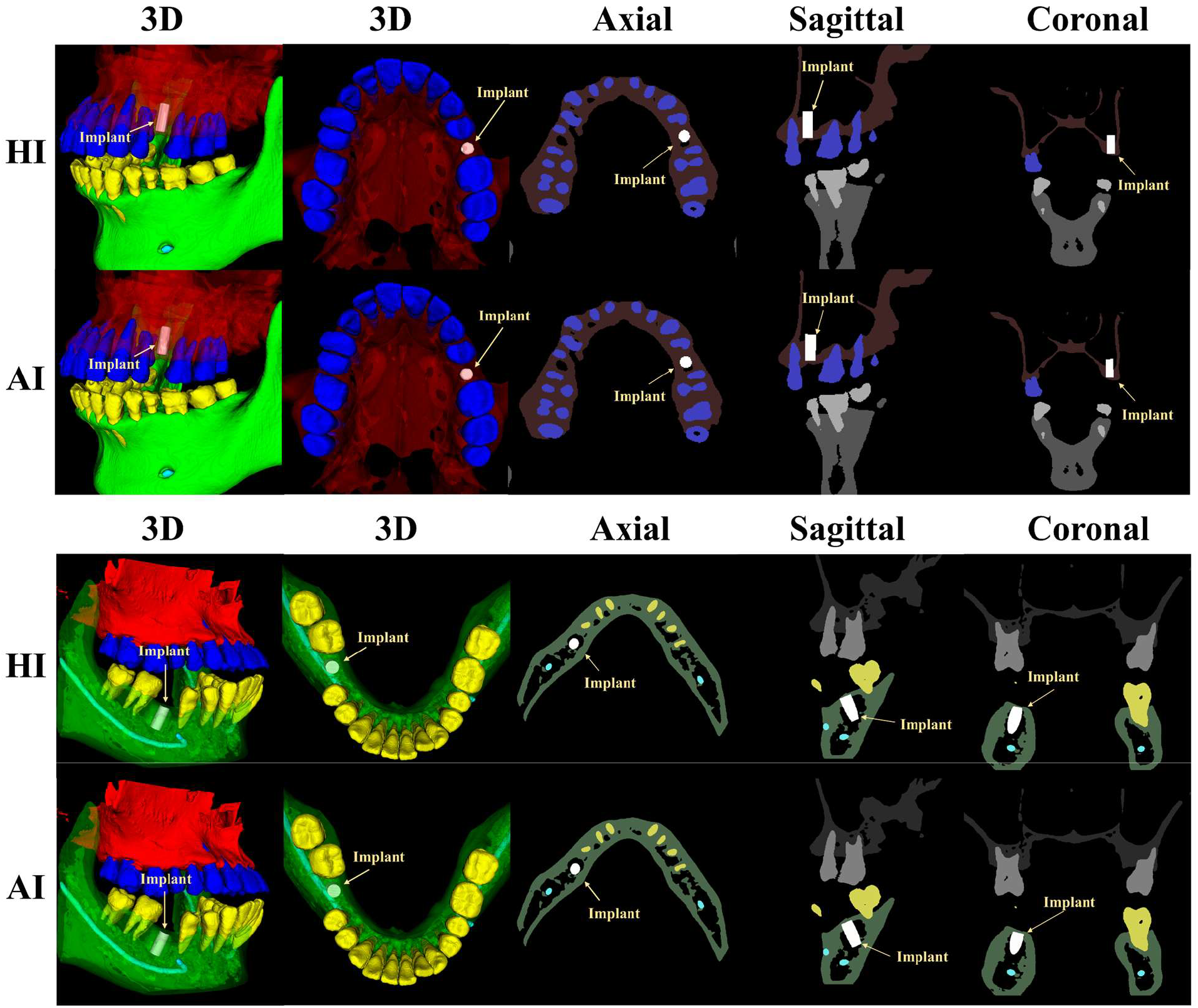
Comparison of AI and HI implant placement positions in single-tooth missing cases (rows 1– 2: maxilla; rows 3–4: mandible). Rows 1 and 3 show the HI plan, while rows 2 and 4 show the AI plan. Red, green, cyan, blue, yellow and white indicate the maxillary bone region, the mandibular bone region, the nerve canal, the maxillary teeth, the mandibular teeth, and the implant respectively.

#### 3.1.2. Quantitative Analysis

We quantified the geometric agreement between the AI and HI plans, as shown in Fig. 6 and Tab. 1. For all single-tooth missing cases, the mean Euclidean centroid distance (CD) was 1.60 ± 1.05 mm. The shoulder deviation (ShD) and apex deviation (AD) were 1.51 ± 1.08 mm and 1.92 ± 1.06 mm, respectively. The axial angle difference (AAD) was 7.16 ± 3.64°. The Dice coefficient (DICE) reached 0.628 ± 0.158, with an average surface distance (ASD) of 0.67 ± 0.39 mm and modified hausdorff distance (MHD) of 0.31 ± 0.30 mm. These metrics, defined in Section 2.7.2, collectively demonstrate a high degree of spatial overlap between the two planning approaches.

**Table 1.** Comparison of implant-level geometric consistency between AI and HI plans for overall, single-tooth, and multiple-tooth missing groups (mean ± SD)

| Metric | Overall (38) | Single (20) | Multiple (18) |
| --- | --- | --- | --- |
| CD (mm) | 1.42 ± 0.80 | 1.60 ± 1.05 | 1.22 ± 0.32 |
| ShD (mm) | 1.38 ± 0.85 | 1.51 ± 1.08 | 1.23 ± 0.48 |
| AD (mm) | 1.71 ± 0.82 | 1.92 ± 1.06 | 1.47 ± 0.30 |
| AAD ($^{\circ}$) | 7.10 ± 3.38 | 7.16 ± 3.64 | 7.05 ± 3.18 |
| DICE | 0.641 ± 0.125 | 0.628 ± 0.158 | 0.656 ± 0.076 |
| MHD (mm) | 0.27 ± 0.23 | 0.31 ± 0.30 | 0.22 ± 0.09 |
| ASD (mm) | 0.60 ± 0.31 | 0.67 ± 0.39 | 0.52 ± 0.16 |

**Fig. 6.**
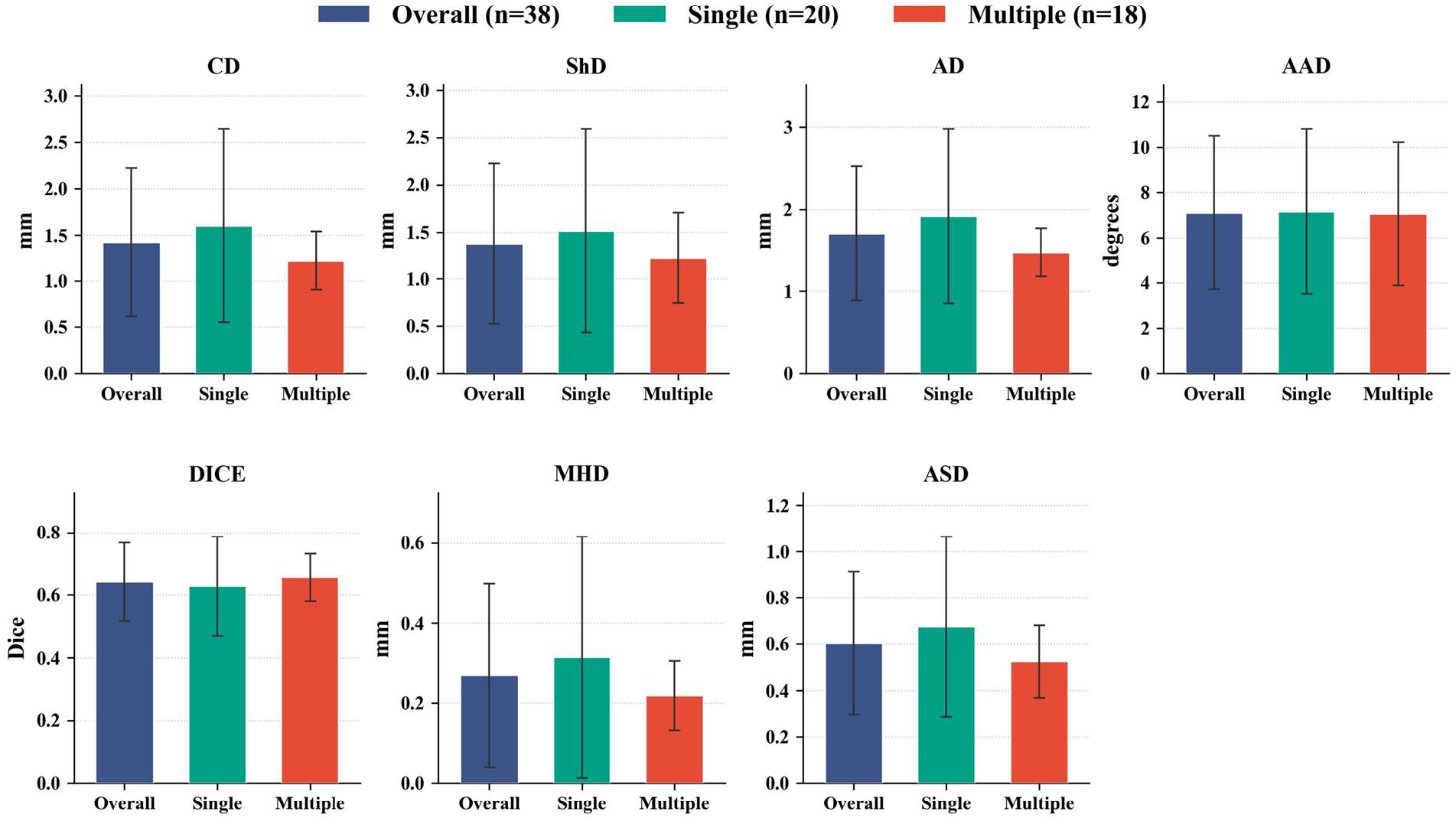
Quantitative comparison of geometric consistency metrics between AI and HI plans across all cases. Metrics include centroid distance, axial angle difference, Dice coefficient, modified Hausdorff distance, and average surface distance, stratified by overall (n=38), single (n=20), and multiple (n=18) groups. Error bars represent standard deviations. Data are shown as mean ± standard deviation (SD).

Clinical safety was evaluated independently of the HI plans using the hard constraints specified in Section 2.7.4. As shown in Fig. 7A, all cases satisfied the clinical safety requirements. The mean minimum distances to the mesial and distal adjacent teeth were 4.20±1.13 mm and 4.22±1.13 mm, respectively, exceeding the required 1.5 mm margin. The minimum distances to the lingual and buccal bone plates were 3.09±1.24 mm and 3.23±1.11 mm, respectively, both exceeding the 1.0 mm and 2.0 mm safety margin. For mandibular cases, the safety distance to the inferior alveolar canal was 4.15±2.06 mm, above the 2.0 mm threshold. These quantitative results confirm that all 20 single-tooth cases achieved full compliance with the predefined safety thresholds.

**Fig. 7.**
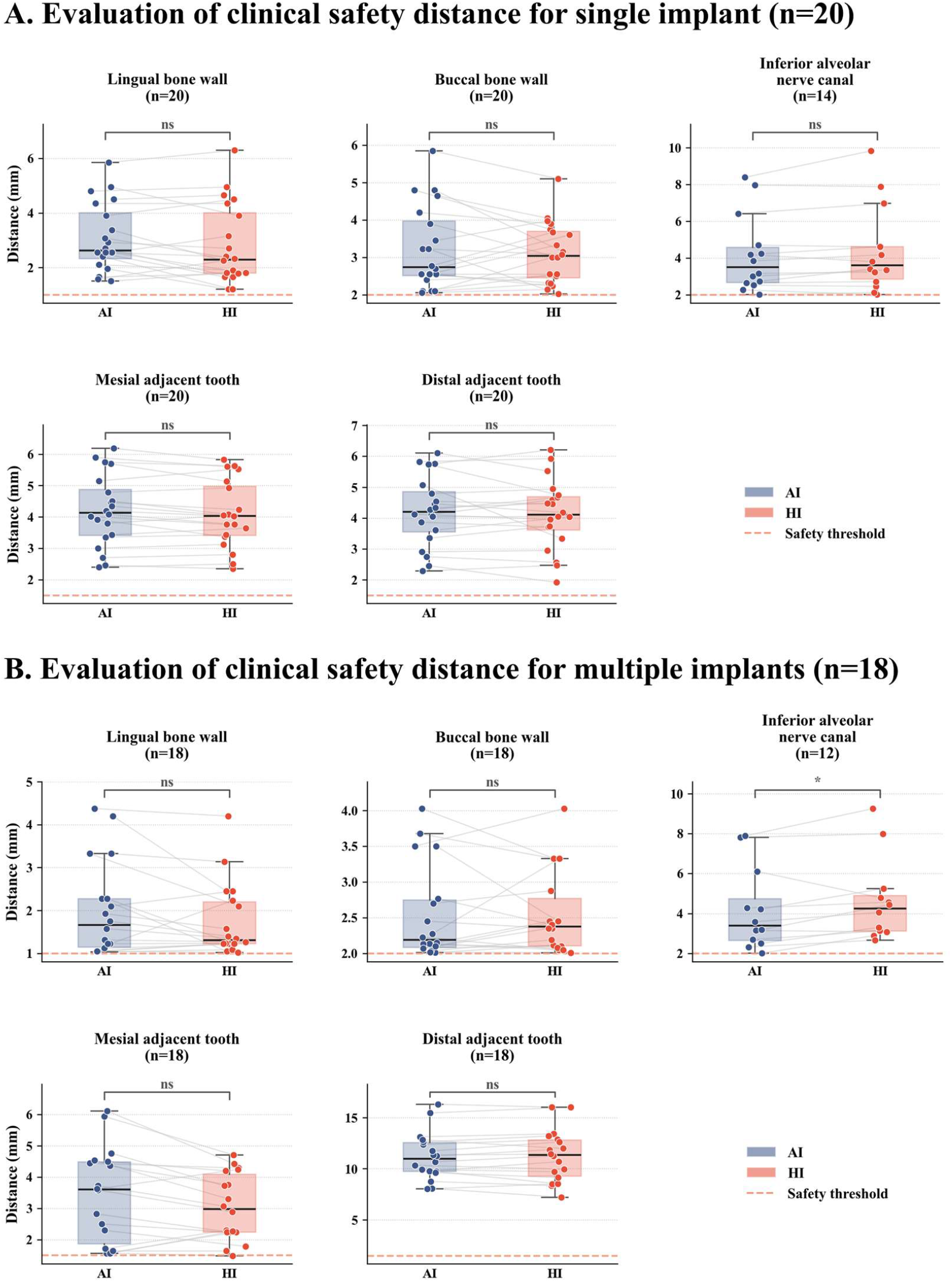
Distribution of safety distances from the planned implants to critical anatomical structures in single-tooth missing cases (part A) and multi-tooth missing cases (part B). This figure presents the measured minimum distances from the planned implants to the lingual bone wall, buccal bone wall, and inferior alveolar nerve for the single implant group (part A, n=20) and the multiple implants group (part B, n=18). The inferior alveolar nerve distances were calculated only for mandibular posterior cases (n=14 for single, n=12 for multiple).

### 3.2. Consecutive Multi-Tooth Implant Planning

#### 3.2.1. Qualitative analysis

At the 3D view (Fig.8), the insertion regions and intra-arch distribution patterns of the implants in the AI plans were generally consistent with the HI plans across the 8 cases. Further comparisons via multi-view 2D cross-sections (Fig. 9, Fig. 10 and Fig. S4) showed that the AI plans achieved good agreement with the HI plans across key anatomical indicators. The distances from the implants to the mandibular canal, adjacent teeth, and bone surfaces were similar, effectively preserving the necessary safety space. The relative positions, spacing, and axial parallelism between the two implants were highly matched with those in the HI plans.Additionally, the buccolingual bone volume allocation showed no obvious tendency to lean toward either side. Overall, the AI plans for multiple-tooth implants maintained good consistency with the HI plans.

**Fig. 8.**
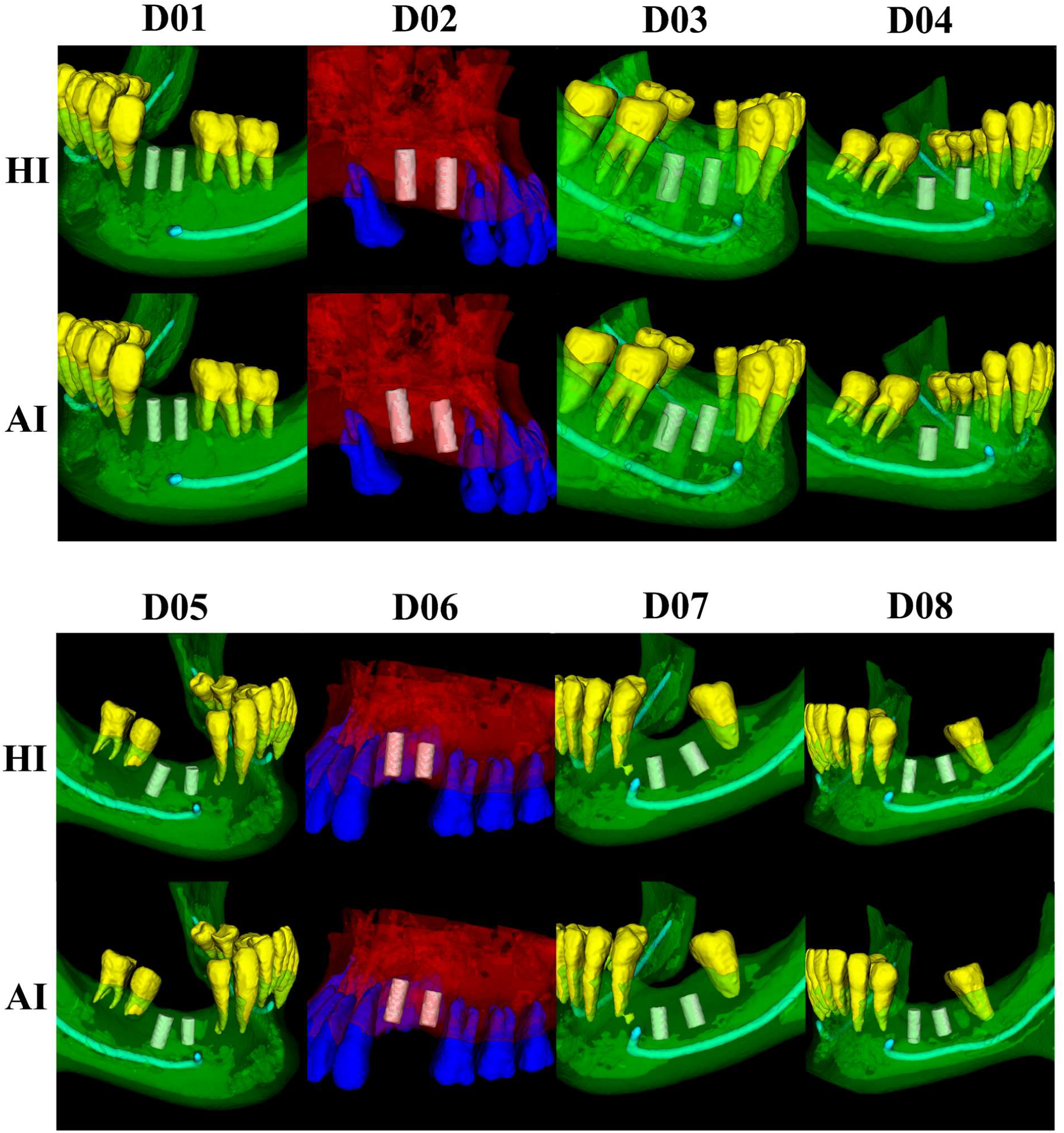
Comparison of AI and HI implant placement positions for eight subjects with consecutive multi-tooth missing. For each subject, the planned implants are displayed within the anatomical context of the jawbone, adjacent teeth, and the mandibular canal, facilitating direct visual comparison between the two planning methods. Red, green, cyan, blue, yellow and white indicate the maxillary bone region, the mandibular bone region, the nerve canal, the maxillary teeth, the mandibular teeth, and the implant, respectively.

**Fig. 9.**
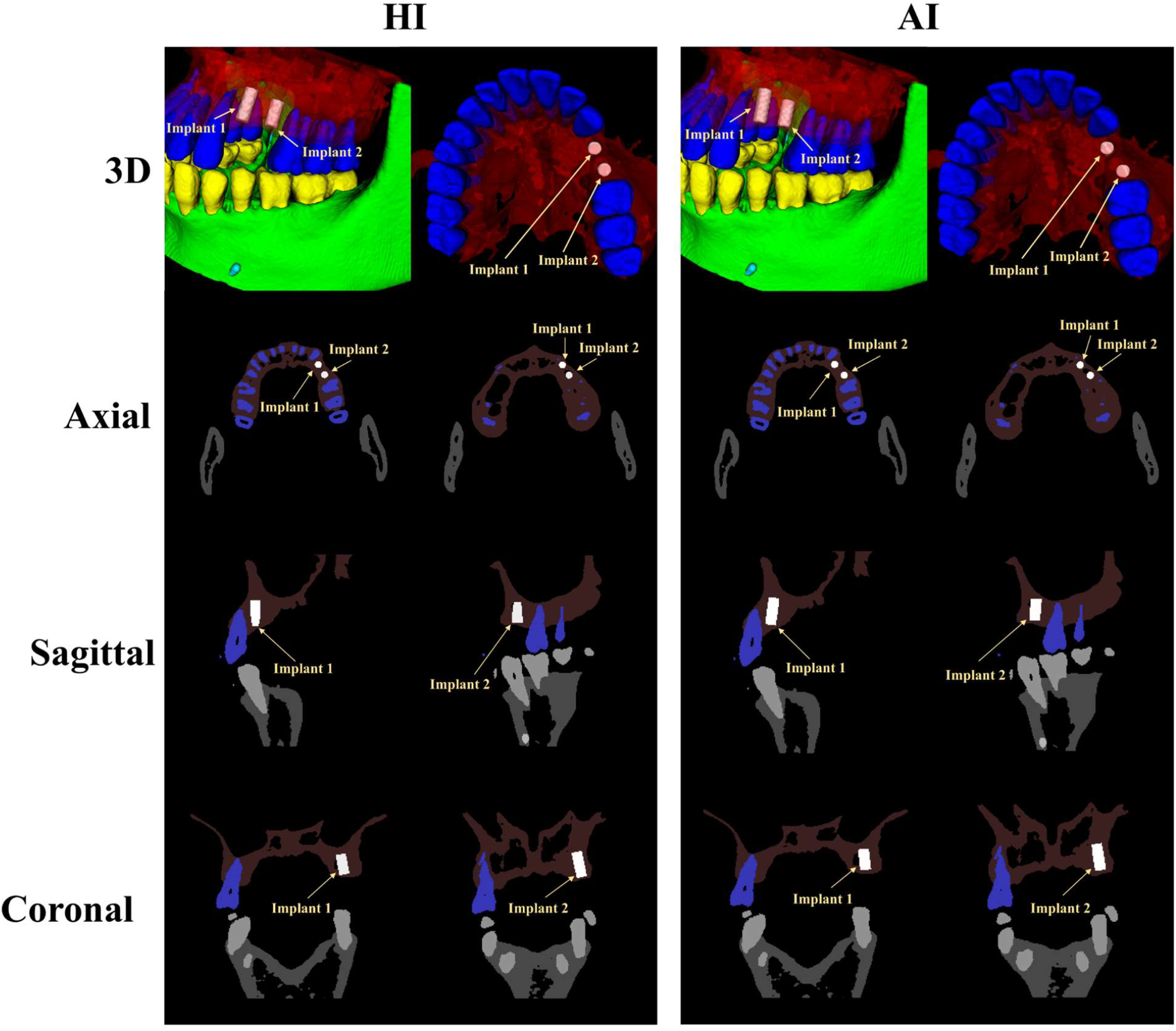
Comparison of AI and HI implant placement positions in a typical maxillary case with two consecutive missing teeth. The left column shows the HI plan, while the right column shows the AI plan. And red, green, cyan, blue, yellow and white indicate the maxillary bone region, the mandibular bone region, the nerve canal, the maxillary teeth, the mandibular teeth, and the implant, respectively.

**Fig. 10.**
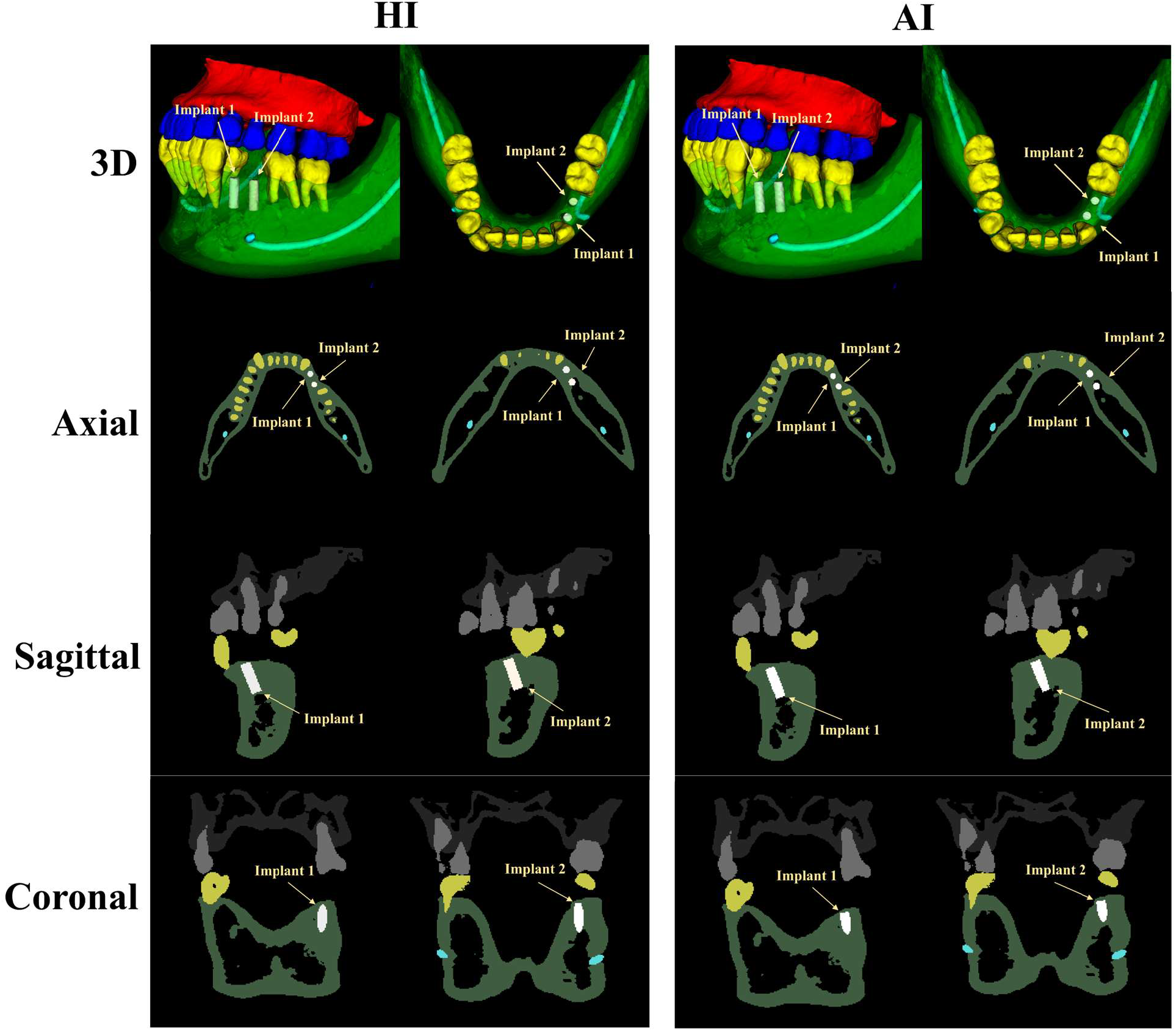
Comparison of AI and HI implant placement positions in a typical mandibular case with two consecutive missing teeth. The left column shows the HI plan, while the right column shows the AI plan. And red, green, cyan, blue, yellow and white indicate the maxillary bone region, the mandibular bone region, the nerve canal, the maxillary teeth, the mandibular teeth, and the implant, respectively.

#### 3.2.2. Quantitative analysis

For multiple missing teeth cases, we evaluated both individual implant accuracy and inter-implant relationships. In terms of individual geometric accuracy (Fig. 6 and Tab. 1), the mean Euclidean centroid distance (CD) between AI and HI plans was 1.22 ± 0.32 mm. The shoulder deviation (ShD) and apex deviation (AD) were 1.23 ± 0.48 mm and 1.47 ± 0.30 mm, respectively. The axial angle difference (AAD) was 7.05 ± 3.18°. The Dice coefficient (DICE) reached 0.656 ± 0.076, with an average surface distance (ASD) of 0.52 ± 0.16 mm and a modified Hausdorff distance (MHD) of 0.22 ± 0.09 mm. These metrics were similar to those obtained in the single-tooth cohort, confirming that the algorithm remains accurate when handling consecutive missing teeth.

For inter-implant relationships, we computed the pairwise metrics defined in Section 2.7.3 (Tab. 2, Supplementary Fig. S1). The absolute differences between AI and HI plans in inter-implant centroid spacing (ICS), shoulder spacing (ISS), and apex spacing (IAS) were 0.92 ± 0.67 mm, 0.81 ± 0.60 mm, and 1.01 ± 0.83 mm, respectively. The inter-implant axial angulation (IAA) deviation was 4.85 ± 2.92°, and the minimum inter-implant surface distance difference was 0.87 ± 0.59 mm. These absolute deviations were small, indicating that the AI framework reproduced the relative configuration of multiple implants with high fidelity to the HI plans.

**Table 2.**
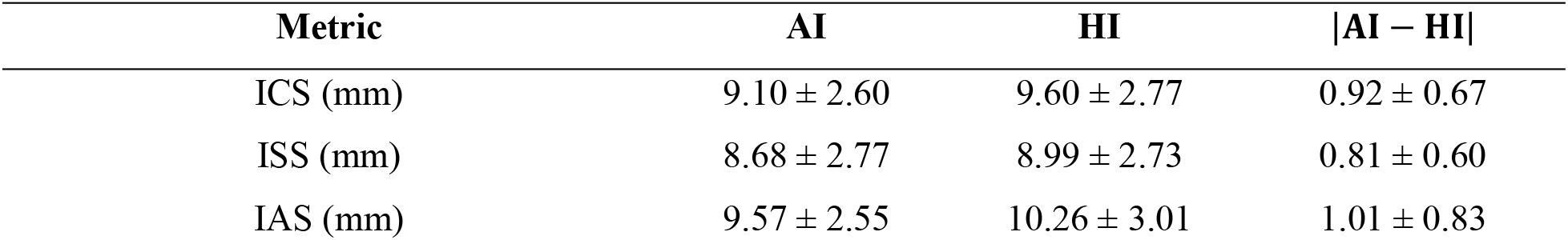
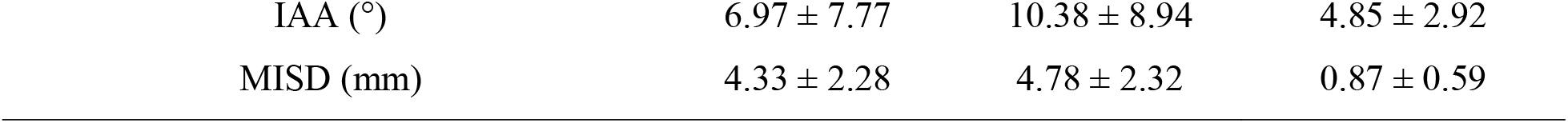
Comparison of inter-implant geometric consistency (spacing, angulation, and surface distance) between AI and HI plans in multiple-tooth implant planning (mean ± SD)

In terms of individual implant safety, all AI plans achieved a 100% compliance rate with the hard constraints for distances to adjacent teeth, buccal and lingual bone plates, and the mandibular canal (Fig. 7B). Collectively, these findings demonstrate that the proposed algorithm not only preserves single-implant accuracy but also maintains robust coordination and safety for multiple implants.

### 3.3. Expert Evaluation

#### 3.3.1. Turing-style Test

A total of 145 forced choice judgments were collected from five expert clinicians across 29 cases. The overall identification accuracy was 52.4% (76 out of 145), with a 95% Wilson confidence interval from 44.3% to 60.4%, and was not significantly different from the 50% chance level (p = 0.62) (Supplementary Fig. S2). Accuracy for single-tooth cases was 48.0% (p = 0.76), and for multi-tooth cases it was 62.2% (p = 0.14), neither reaching statistical significance. Individual expert accuracy ranged from 34.5% to 62.1%, all within the range of random variation. Confidence scores were generally high, with most responses rating 4 or 5, and the mean confidence was nearly identical between correct and incorrect judgments, at approximately 3.97 and 4.01, indicating pronounced overconfidence. The row normalized confusion matrix showed a balanced error distribution, and the case-level ROC AUC was 0.476, closely aligning with the chance line. Inter rater reliability, assessed by Fleiss’ kappa, was approximately 0.06, indicating negligible agreement among experts. These results demonstrate that under blinded conditions, experts could not reliably distinguish AI plans from HI plans, confirming that the proposed framework passes the Turing style discrimination test.

#### 3.3.2. Expert Clinical Rating (ECR)

Five expert clinicians rated the 29 AI plans using a 5-point Likert scale, the results were represented in Supplementary Fig. S3. The overall mean score was 4.32 ± 0.71, with a median of 4 and scores ranging from 2 to 5. For single-tooth cases, the mean score was 4.44 ± 0.64, with 52% of ratings at 5 and 40% at 4, and almost no ratings below 3. For multi-tooth cases, the mean score was 4.04 ± 0.80, with 31% at 5 and 44% at 4, and only 2% at 2. Across both groups, the vast majority of ratings fell in the 4 to 5 range, indicating high clinical acceptability. Individual rater means ranged from 3.93 to 4.45, with one rater being systematically more conservative. A Friedman test revealed a significant rater effect (χ^2^ = 15.67, p = 0.0035), and pairwise Wilcoxon tests confirmed that the conservative rater differed significantly from three others. Pairwise quadratic weighted kappa values averaged around 0.33, reflecting moderate inter rater agreement with notable variability. The intraclass correlation coefficient for the average of five raters, ICC(C,k), was 0.717 with a bootstrap 95% confidence interval from 0.481 to 0.840, which falls in the good reliability range. Overall, the expert clinical ratings indicate that the AI plans were of high quality and clinically acceptable.

## 4. Discussion

### 4.1 Key findings and clinical relevance

This study proposed an interpretable framework for automated implant planning and validated it on 29 clinical cases, including 20 single-tooth and 9 consecutive multi-tooth edentulous sites. The AI plans showed high geometric agreement with human expert plans. For single-tooth cases, the centroid distance (CD) was 1.60±1.05 mm, and for multi-tooth cases it was 1.22±0.32 mm. The axial angle difference (AAD) was approximately 7° in both groups, while the average surface distance (ASD) and modified Hausdorff distance (MHD) remained small. The Dice coefficient ranged from 0.63 to 0.66. This relatively modest overlap is largely explained by the small volume of dental implants, where a positional deviation of 1–2 mm can markedly reduce volumetric overlap. In this context, ASD, MHD, and expert ratings provide more clinically meaningful indicators of agreement than Dice alone. For multi-tooth cases, the absolute differences between AI and HI plans in inter-implant centroid spacing, shoulder spacing, and apex spacing were all below approximately 1.1 mm. The inter-implant axial angulation differed by 4.85±2.92°, and the minimum inter-implant surface distance differed by 0.87±0.59 mm. These results indicate that the AI framework reproduced the relative configuration defined by human experts. More importantly, all AI plans achieved 100% compliance with the predefined safety thresholds, including minimum distances to adjacent teeth, buccal and lingual bone walls, and the mandibular canal. This highlights a clear advantage of the proposed framework in prioritizing safety.

In terms of clinical acceptability, five experts rated the 29 AI plans with a mean score of 4.32±0.71, with most ratings concentrated in the 4–5 range. The intraclass correlation coefficient, ICC(C,k), was 0.717, indicating good reliability. In the Turing-style discrimination test, the overall accuracy of experts in distinguishing AI from HI plans was 52.4% (95% Wilson CI: 44.3%–60.4%), which was not significantly different from the 50% chance level (p=0.62). Accuracy was 48.0% for single-tooth cases and 62.2% for multi-tooth cases, neither reaching statistical significance. Although experts generally reported high confidence in their judgments, their discrimination accuracy remained close to chance. Under blinded conditions, therefore, AI and HI plans were difficult to distinguish visually and clinically, supporting the clinical credibility of the proposed framework. Overall, the framework generates safe, interpretable, and clinically acceptable implant plans. It is intended as a decision-support tool that improves planning consistency and efficiency, rather than a replacement for human experts.

### 4.2. Hybrid Distance Measurement Strategy

The main advantage of this work lies in separating perception from reasoning. The deep network is responsible only for CBCT anatomical segmentation, while the planning logic is implemented through explicit geometric modeling and constrained optimization. Compared with end-to-end deep learning methods that directly regress implant parameters [11-13, 16] or treat implants as segmentation labels [10], the proposed framework provides a complete audit trail: dental arch extraction, ROI cropping, axis initialization, distance measurement, and position optimization.

Each step corresponds to an operation that clinicians can understand. The implant axis is obtained by spherical linear interpolation (SLERP) between the long axes of adjacent teeth, analogous to the clinical concept of following the emergence profile. The optimization moves the implant along the arch tangent, buccolingual normal, and implant axis, rather than in arbitrary Cartesian directions. This design improves both interpretability and search efficiency. The lexicographic optimization further reflects clinical priorities: hard constraints on bone depth, nerve distance, and adjacent teeth are satisfied first, followed by minimization of mesiodistal imbalance, and finally buccolingual bone balance. As a result, the final plan remains safe even when perfect balance cannot be achieved.

For distance measurement, this study adopts a hybrid strategy combining ray casting and distance transform. Ray casting is well suited for continuous bone surfaces. A backward-tracking procedure avoids mistaking internal voids or segmentation defects for the true cortical boundary. Distance transform is suitable for discrete structures such as adjacent teeth and the mandibular canal, allowing rapid queries of minimum distances during iterative optimization. This combination balances robustness and computational efficiency. Compared with existing end-to-end methods, the proposed framework differs in three main respects. First, it does not rely on large amounts of expert-annotated implant plans. The deep network requires only segmentation labels, while the planning logic is encoded as explicit rules and constraints, substantially reducing dependence on implant planning annotations. Second, explicit hard constraints guarantee safety margins at inference time, rather than relying on loss-function penalties. Third, the planning logic is based on geometric constraints and clinical priors rather than a specific training distribution, allowing a unified framework to handle both single-tooth and consecutive multi-tooth cases. Compared with conventional manual planning, the proposed method can improve planning consistency, reduce inter-clinician variability and planning time, while preserving a traceable and modifiable geometric reasoning process. For multi-tooth cases, a global coordination strategy is introduced, including parallelism constraints, inter-implant distance constraints, a common insertion path constraint, and an arch-distribution prior, to ensure prosthetic feasibility and long-term stability.

### 4.3. Limitations and future work

Despite these promising results, several limitations remain. First, the current optimization considers only geometric constraints and does not yet integrate biomechanical factors such as bone density distribution, occlusal load direction, or finite element analysis. Future work could incorporate biomechanical models to make planning more consistent with physiological and mechanical requirements[23]. Second, the initialization is still based on the midpoint of adjacent teeth and a SLERP-derived axis, which is relatively rule-based. A deep learning model could be used to predict a rough initial implant pose, which would then be refined by the geometric optimizer, combining data-driven pattern recognition with safety and interpretability. Third, the multi-tooth validation included only 9 cases, most of which involved two consecutive missing teeth. The common insertion path constraint, which becomes active only when three or more implants are involved, has not been sufficiently validated. Future studies should include multi-center, prospective, and larger cohorts, covering longer edentulous spans, full edentulism, different ethnic groups, CBCT protocols, and implant systems, and should report long-term implant survival, osseointegration, esthetic outcomes, and patient satisfaction. Addressing these directions will advance the system toward a robust, clinically deployable tool[24].

## 5. Conclusion

In summary, this work shows that a geometry-driven, interpretable planning framework can generate implant positions that closely match expert plans while strictly satisfying clinical safety constraints. By limiting deep learning to anatomical segmentation and encoding the planning logic as explicit geometric operations, the approach offers a transparent alternative to end-to-end black-box models. Its unified formulation for both single and consecutive multiple missing teeth further supports its potential for broader clinical use. Although larger multi-center studies and biomechanical extensions are still needed, the results suggest that this framework can serve as a reliable decision-support tool, helping clinicians improve planning consistency and efficiency without giving up control over the final plan.

## Data Availability

The data supporting the findings of this study are not publicly available due to patient privacy and institutional and ethical restrictions.

