## Supplementary Materials for "A Unified and Interpretable Geometric Optimization Framework for Intelligent Automated Implant Planning in CBCT from Single to Consecutive Multiple Missing Teeth"

Funding: This work was supported by the Early-Career Young Scientists and Technologies Project of Jiangxi Province (No. 20252BEJ730261); the 2025 Research Startup Fund for Full-time Introduced Talents in Natural Sciences (Project No. 2025YJRC0015); and the National Natural Science Foundation of China Youth Science Fund Project (Project No. 82401196).

**Conflicts of Interest**: None of the authors has a financial interest in any of the products, devices, or drugs mentioned in this manuscript.

**Manuscript information: 39 text pages, 10 figures, 2 tables.**

**(Supplementary Information: 9 text pages, 5 figures)**

**SI-1. Inclusion Criteria**

The inclusion criteria were as follows: 1) age ≥ 18 years; 2) presence of at least one single-tooth missing site or a consecutive edentulous span of 2–3 adjacent teeth in the maxilla or mandible; 3) adequate bone volume for implant placement without the need for major bone augmentation; and 4) CBCT images with sufficient quality for anatomical identification. Exclusion criteria were: 1) CBCT images with severe metal artifacts, motion artifacts, or poor image quality compromising diagnostic assessment; 2) edentulous sites with adjacent teeth indicated for extraction (e.g., severe periodontal bone loss or extensive caries); 3) radiographic evidence of pathological conditions in the jawbone (e.g., cysts, tumors); and 4) incomplete healing of extraction sockets.

**SI-2. Ground Truth Implant Planning**

For each edentulous site, a senior oral surgeon performed manual virtual implant planning using the implant planning module in 3D Slicer, following a restoration-driven protocol. The planning process considered three core clinical requirements: restoration orientation (aligning the implant axis with the anticipated prosthetic crown), bone adaptation (selecting appropriate implant dimensions and position to maximize cortical bone engagement), and anatomical safety (maintaining safe distances from adjacent teeth, buccal/lingual bone walls, and the mandibular nerve canal). The planned implants were selected from a clinically available set of standard dimensions, including diameters of 3.3, 4.1, and 4.8 mm and lengths of 8 and 10 mm, reflecting common clinical practice. The HI plans served as the ground truth for evaluating the automated planning method. The quality of all ground truth plans was independently assessed by five experienced implantologists using a 5-point Likert scale (1 = clinically unacceptable, 5 = clinically excellent), with only plans scoring ≥ 4 included in the final dataset.

**Table S1. Scoring criteria for single implant.**

| Score | Evaluation Criteria |
|---|---|
| 5 points | No modification required; implant size, direction, and three-dimensional position are all satisfactory. |
| 4 points | Clinically acceptable; one of size, direction, or three-dimensional position requires minor adjustment. |

| | |
|---|---|
| 3 points | Clinically acceptable; two of size, direction, or three-dimensional position require minor adjustment. |
| 2 points | Clinically acceptable; all three (size, direction, and three-dimensional position) require minor adjustment. |
| 1 point | Major revision required; one or more items are clearly inaccurate, and the case is clinically unacceptable (e.g., damage to the inferior alveolar nerve). |

**Table S2. Scoring criteria for dual implants.**

| Score | Evaluation Criteria |
|---|---|
| 5 points | No modification required. The size, direction, and three-dimensional position of each of the two implants are satisfactory; and the inter-implant positional relationships meet the following: inter-implant distance ⩾3 mm, height difference at the restorative platform <1 mm, and distance to adjacent tooth root ⩾1.5 mm; if in the mandibular posterior region, the distance from each implant to the mandibular canal is ⩾2 mm. |
| 4 points | Clinically acceptable but requiring minor adjustment. Meets one of the following: ① one of the individual implant factors (size, direction, or 3D position) requires minor adjustment; ② only one of the three inter-implant positional items (e.g., inter-implant distance 2.5–3 mm) shows minor deviation, but does not compromise the final restoration; ③ in the mandibular posterior region, the implant-to-canal distance is 1.5–2 mm, with no risk of nerve injury. |
| 3 points | Clinically acceptable but requiring minor adjustment. Meets one of the following: ① two of the individual implant factors require minor adjustment; ② two of the three inter-implant positional items show minor deviation; ③ one individual implant factor plus one inter-implant positional item show minor deviation; ④ in the |

| | |
|---|---|
| | mandibular posterior region, the implant-to-canal distance is 1.5–2 mm, combined with a minor deviation in one individual implant factor or one inter-implant positional item. |
| 2 points | Clinically acceptable but requiring minor adjustment. Meets one of the following: ① all three individual implant factors of a single implant require minor adjustment; ② all three inter-implant positional items show minor deviation; ③ two individual implant factors plus one inter-implant positional item show minor deviation; ④ one individual implant factor plus two inter-implant positional items show minor deviation; ⑤ in the mandibular posterior region, the implant-to-canal distance is 1.5–2 mm, combined with minor deviation in two individual implant factors or two inter-implant positional items. |
| 1 point | Major revision or re-planning required. Meets any of the following: ① any single implant has a significant inaccuracy that is clinically unacceptable (e.g., impingement on the inferior alveolar nerve, perforation of cortical bone, etc.); ② the inter-implant positional relationships are severely substandard (e.g., inter-implant distance <2 mm, or severe height mismatch preventing a common path of insertion); ③ in the mandibular posterior region, the implant-to-canal distance is <1.5 mm, with a definite high risk of nerve injury. |

**Table S3. Scoring criteria for multiple implants. （>2）**

| Score | Evaluation Criteria |
|---|---|
| 5 points | No modification required. The size, direction, and 3D position of all implants are satisfactory; and their mutual positional relationships meet: inter-implant spacing ⩾3 mm, restorative platform height difference <1 mm, distance to adjacent tooth root ⩾1.5 mm; if in |

| | |
|---|---|
| | the posterior mandible, the distance from each implant to the mandibular canal is ≥2 mm. |
| 4 points | Clinically acceptable, but minor adjustments needed. Meets one of the following: ① one single implant factor requires minor adjustment; ② only one of the three mutual positional items (e.g., inter-implant spacing 2.5–3 mm) shows minor deviation, but does not affect the final restoration; ③ in the posterior mandible, the implant-to-mandibular canal distance is 1.5–2 mm, with no risk of nerve injury. |
| 3 points | Clinically acceptable, but minor adjustments needed. Meets one of the following: ① two single implant factors require minor adjustment; ② two of the three mutual positional items show minor deviation; ③ one single implant factor plus one mutual positional item show minor deviation; ④ in the posterior mandible, the implant-to-mandibular canal distance is 1.5–2 mm, combined with one minor deviation in either a single implant factor or a mutual positional item. |
| 2 points | Clinically acceptable, but minor adjustments needed. Meets one of the following: ① all three single implant factors of a single implant require minor adjustment; ② all three mutual positional items show minor deviation; ③ two single implant factors plus one mutual positional item show minor deviation; ④ one single implant factor plus two mutual positional items show minor deviation; ⑤ in the posterior mandible, the implant-to-mandibular canal distance is 1.5–2 mm, combined with either two single implant factors or two mutual positional deviations. |

| | |
|---|---|
| 1 point | Major correction or re-planning required. Meets any one of the following: ① any single implant has a significant inaccuracy that is clinically unacceptable (e.g., impingement on the inferior alveolar nerve, perforation of the cortical bone, etc.); ② the mutual positional relationships seriously fail to meet standards (e.g., inter-implant spacing <2 mm, or severe height mismatch making restoration impossible); ③ in the posterior mandible, the implant-to-mandibular canal distance is <1.5 mm, with clear risk of nerve injury. |

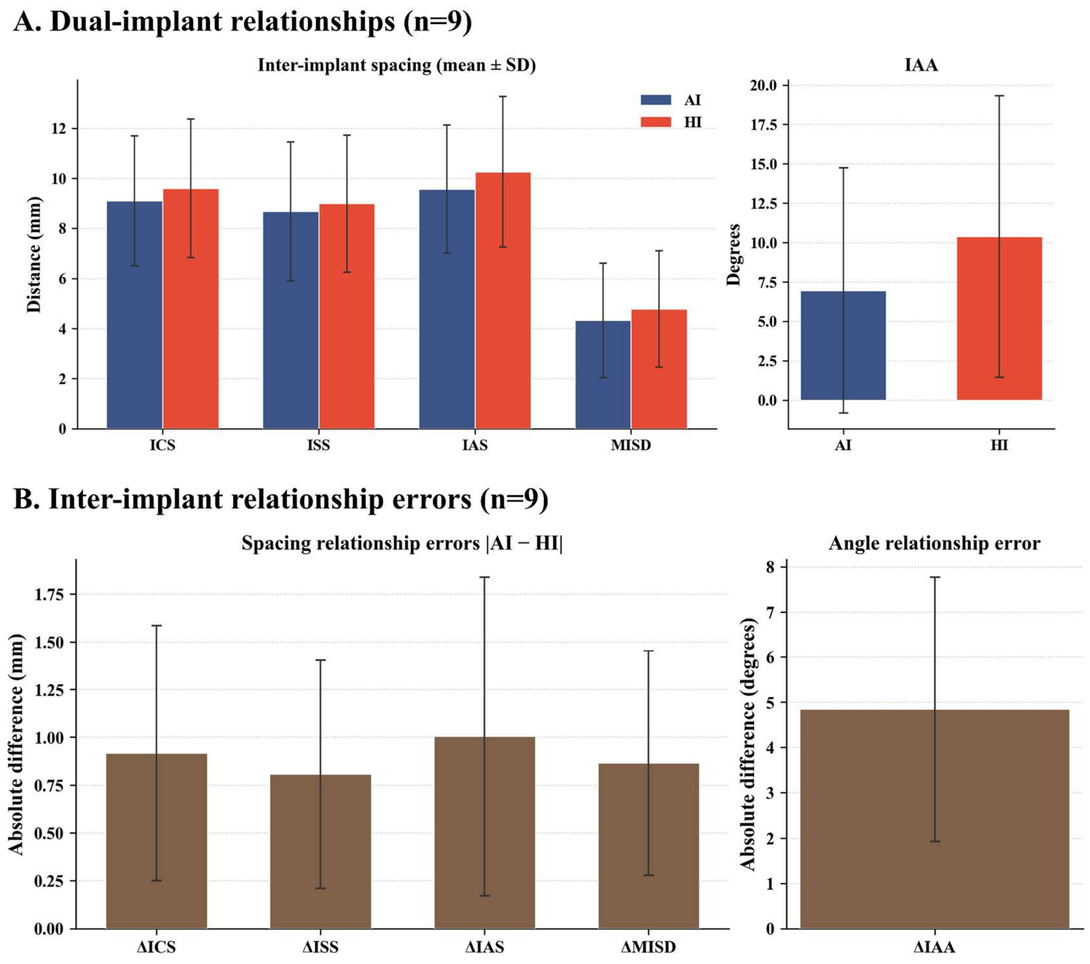


**Figure S1. Comparison of inter-implant relationships between AI and HI plans (n=9).** (A) Mean (± SD) spacing metrics and axial angle for AI (blue) and HI (red) implants.

(B) Absolute pairwise errors ($|AI - HI|$) for the corresponding metrics.

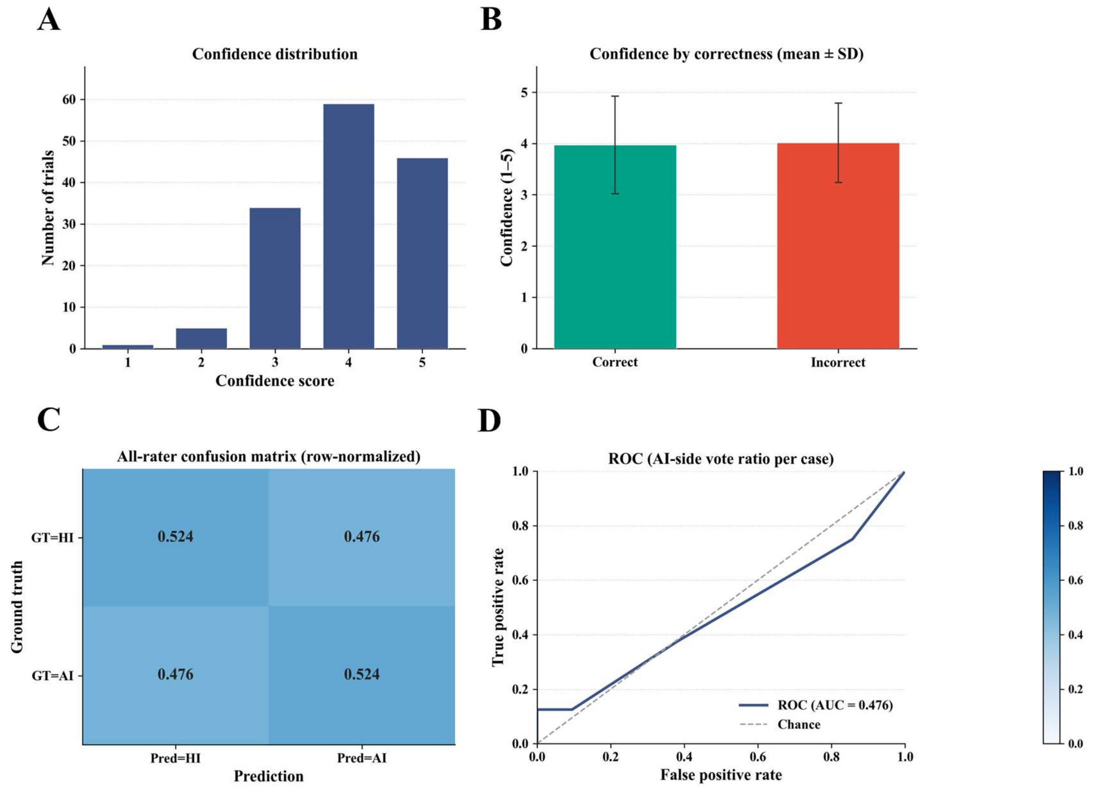


**Fig. S2. Confidence distribution, confusion matrix, and ROC curve analysis for the Turing-style discrimination test.** (A) Confidence distribution: The bar chart shows the distribution of confidence scores assigned by experts to their own judgments across 145 trials. The x-axis represents the confidence score (1–5), and the y-axis represents the corresponding number of trials. (B) Confidence by correctness (mean ± SD): The bar chart groups the experts' judgments into "correct" and "incorrect" categories, and displays the mean confidence scores for each group. The x-axis is divided into correct and incorrect groups, and the y-axis indicates the mean confidence score. Data are shown as mean ± standard deviation (SD). (C) All-rater confusion matrix (row-normalized): Rows represent the ground-truth labels (GT=HI or GT=AI), columns represent the predicted labels (Pred=HI or Pred=AI), and the cell values indicate the proportions of predictions. (D) ROC (AI-side vote ratio per case): The y-axis is the true positive rate, and the x-axis is the false positive rate. The ROC curve (solid line) and the chance baseline (dashed line) are plotted, with the AUC value annotated.

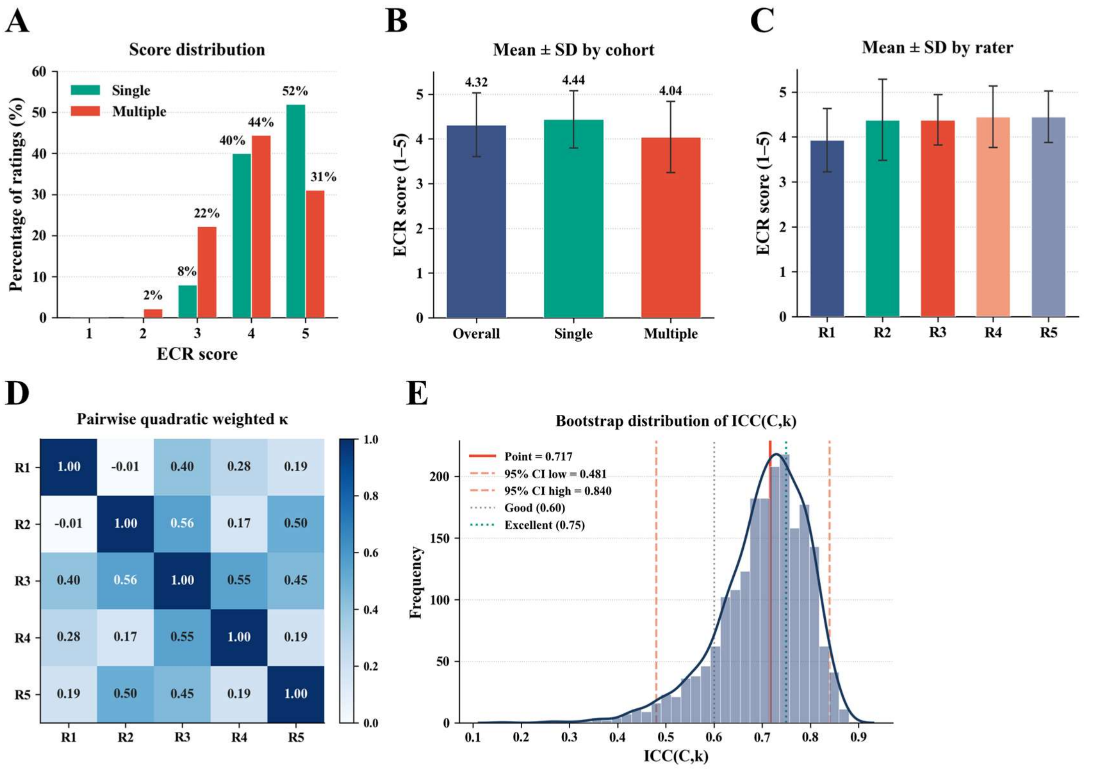


| | R1 | R2 | R3 | R4 | R5 |
|---|---|---|---|---|---|
| R1 | 1.00 | -0.01 | 0.40 | 0.28 | 0.19 |
| R2 | -0.01 | 1.00 | 0.56 | 0.17 | 0.50 |
| R3 | 0.40 | 0.56 | 1.00 | 0.55 | 0.45 |
| R4 | 0.28 | 0.17 | 0.55 | 1.00 | 0.19 |
| R5 | 0.19 | 0.50 | 0.45 | 0.19 | 1.00 |

**Fig. S3. Expert Clinical Rating (ECR) results for AI implants .**This figure summarizes the distribution of ECR scores, mean scores across cohorts and raters, inter-rater agreement, and the reliability of the averaged ratings.(A) The bar chart shows the percentage of ratings for each ECR score (1–5), separately for single-tooth and multiple-tooth cases; the x-axis denotes the score level and the y-axis denotes the percentage of ratings.(B) The bar chart displays the mean ECR score with standard deviation for all cases combined, single-tooth cases, and multiple-tooth cases; the x-axis indicates the cohort and the y-axis indicates the mean score. Data are shown as mean ± standard deviation (SD). (C) The bar chart presents the mean ECR score with standard deviation for each of the five expert raters (R1–R5); the x-axis labels the rater and the y-axis gives the mean score. Data are shown as mean ± standard deviation (SD). (D) The matrix reports the pairwise quadratic weighted kappa coefficients among the five raters; the rows and columns correspond to the raters, and each cell value represents the kappa coefficient for that pair.(E) The histogram illustrates the bootstrap distribution of the intraclass correlation coefficient for the average of five raters (ICC(C, k)); the x-axis is the ICC value, the y-axis is the bootstrap frequency, and the vertical lines mark the point estimate, the 95% confidence interval bounds, and reference thresholds for good (0.60) and excellent (0.75) reliability.

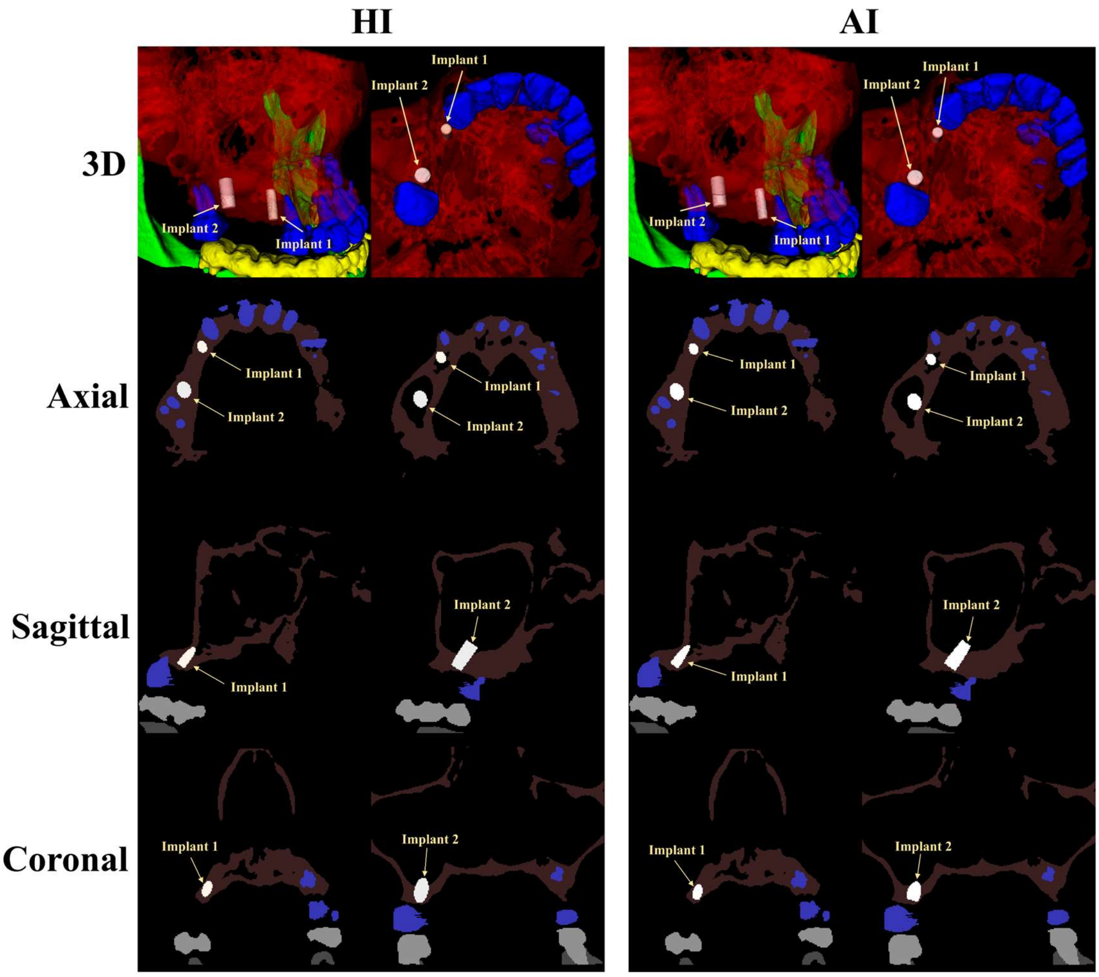


**Fig. S4. Comparison of AI and HI implant placement positions in a typical maxillary case with three consecutive missing teeth.** In the final output, only the two terminal implants are retained for n>2. The left column illustrates the HI plan, while the right column presents the AI plan. And red, green, cyan, blue, yellow and white indicate the maxillary bone region, the mandibular bone region, the nerve canal, the maxillary teeth, the mandibular teeth, and the implant, respectively.

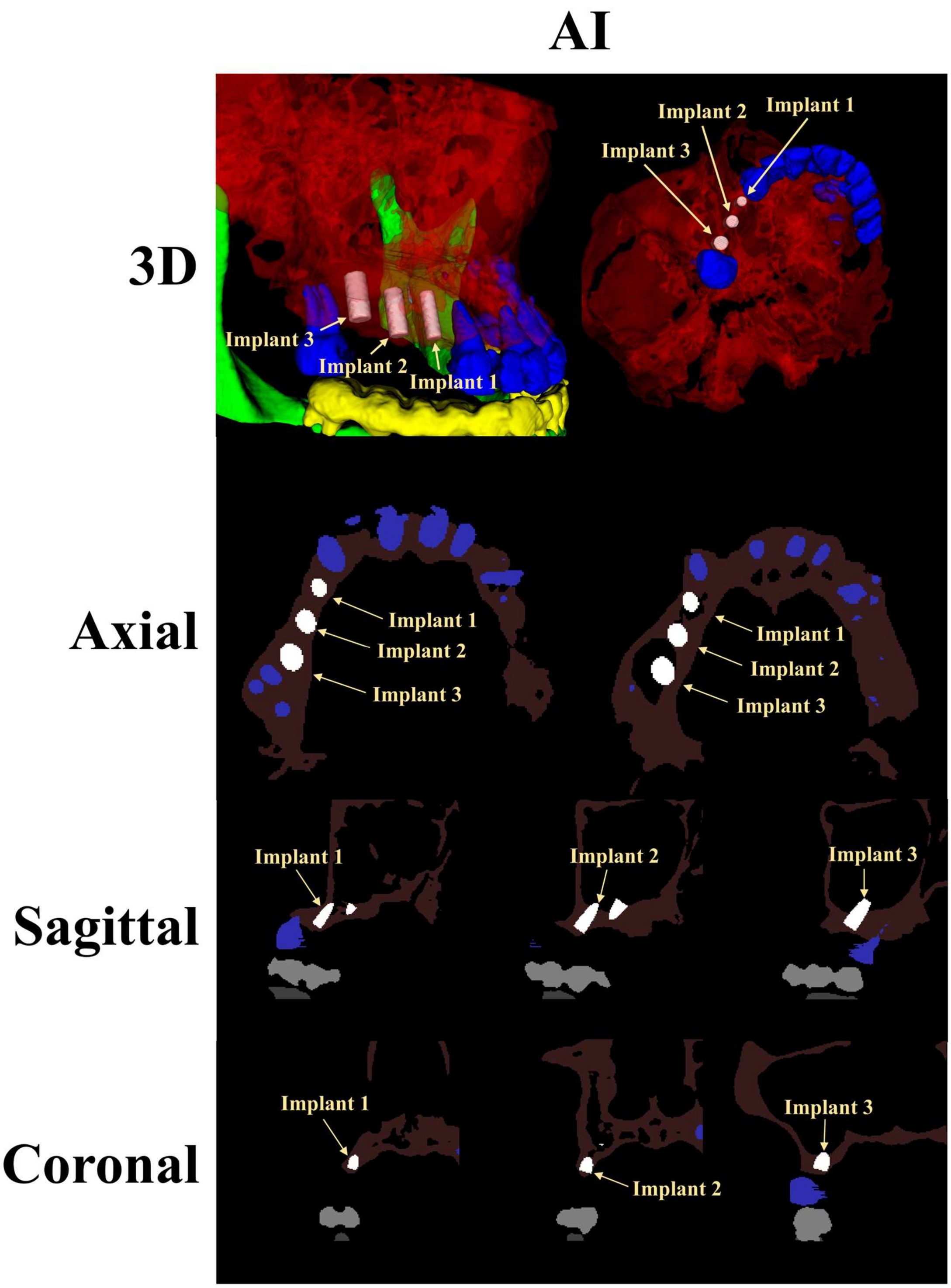


**Fig. S5. Initialization stage of the AI plan for the same maxillary case with three consecutive missing teeth as presented in Fig. S4.** At this stage, three implant positions (Implant 1, Implant 2, and Implant 3) are initialized. The rows from top to bottom show the 3D view, axial, sagittal, and coronal cross-sections, respectively. Red, green, cyan, blue, yellow and white indicate the maxillary bone region, the mandibular bone region, the nerve canal, the maxillary teeth, the mandibular teeth, and the implant, respectively.
